# Tandem repeat expansions in *DAPK1*, *ANK3*, and *RPL14* are associated with diverse neurodegenerative diseases

**DOI:** 10.64898/2026.08.06.26358503

**Authors:** Gabrielle N. Altman, Bharati Jadhav, Paras Garg, Mariya Shadrina, Celine A. Manigbas, William Lee, Shrishtee Kandoi, Alejandro Martin-Trujillo, Andrew J. Sharp

## Abstract

Tandem repeat expansions (TREs) cause over 50 neurological conditions, yet their contribution to neurodegenerative disease risk at a population scale remains incompletely characterized. We performed a TRE association study across 6,539 short tandem repeat loci in 276,411 individuals from the UK Biobank and 44,370 individuals from the All of Us Research Program, using two composite neurodegenerative phenotypes to increase statistical power and capture pleiotropic effects. Meta-analysis across the two cohorts identified associations at eight established pathogenic TRE loci, including *C9orf72*, *DMPK*, *HTT*, *ATXN2*, *ATXN3*, *CACNA1A*, *CNBP*, and *PPP2R2B*, recovering known disease-associated expansions from short-read sequencing data at biobank scale. We also identified candidate associations at three additional loci. An intronic AATAA expansion in *DAPK1* reached significance (q = 0.0045), with fine-mapping and conditional analysis supporting the repeat as the likely variant underlying the association. An intronic ATTTT expansion in *ANK3* (q = 0.034) was observed exclusively in individuals of African and Latino/admixed American ancestry, underscoring the importance of ancestrally diverse cohorts for genetic discovery. An exonic polyalanine expansion in *RPL14* was also significant (q = 0.039), where longer alleles were consistently associated with reduced *RPL14* expression across independent datasets. Together, these findings identify candidate risk loci for neurodegenerative disease that may expand the contribution of TREs to neurodegenerative disease beyond known repeat expansion disorders.

## Introduction

Genome-wide association studies (GWAS) have successfully identified common single nucleotide variants (SNVs) associated with a variety of neurodegenerative diseases.^1–3^ Rare coding variants, copy number variants, and structural variants have also contributed to our understanding of the genetic architecture of neurodegenerative diseases, but still a substantial proportion of phenotypic heritability remains unexplained.^4^

Tandem repeat expansions (TREs) result from an increase in the number of copies of a short repetitive DNA sequence, or short tandem repeat (STR), beyond a normal range. They are known to cause over 50 neurological conditions, including neurodegenerative diseases such as Huntington’s disease, spinocerebellar ataxias, frontotemporal dementia (FTD), and amyotrophic lateral sclerosis (ALS).^5^ The pathogenic repeat length threshold varies substantially across loci, and even within a given locus, different expansion lengths can lead to distinct clinical presentations. This is seen in *ATXN2*, where intermediate-length expansions (27-33 repeats) of a coding CAG repeat increase ALS risk, while full expansions (≥33 repeats) cause spinocerebellar ataxia (SCA) type 2, and CAA-interrupted expansions of similar length have been associated with parkinsonism rather than ataxia.^6,7^

Pleiotropy across neurodegenerative diseases is well established for both SNVs and TREs. Cross-phenotype GWAS studies have identified shared genetic risk loci across Alzheimer’s disease (AD), Parkinson’s disease (PD), ALS, and FTD, and there are modest genetic correlations between various combinations of neurodegenerative diseases.^3,4,8^ At the TRE level, large expansions of a GGGGCC intronic repeat in *C9orf72* can cause both ALS and FTD, with some carriers developing features of both.^9^ Clinical overlap further complicates single-disease analyses, as *C9orf72* expansion carriers presenting with memory impairment or parkinsonism may be misdiagnosed as AD or PD, leading to their exclusion from FTD or ALS association studies,^9^ and *C9orf72* expansions are found in clinically diagnosed AD cases.^10^ Combining overlapping neurodegenerative diagnoses into a composite phenotype addresses both problems. It recovers misclassified cases that would otherwise be excluded from single-disease analyses and increases statistical power to detect pleiotropic loci with shared effects across diseases.

Until recently, TREs have been difficult to study at large scale due to challenges in genotyping repetitive sequences in short-read data, especially when the repeat length is close to or exceeds the read length used in sequencing.^11^ Recent advances in bioinformatics tools such as ExpansionHunter,^12^ combined with the increased availability of large biobank cohorts with short-read genome sequencing (GS) data and linked Electronic Health Record (EHR) data, such as the UK Biobank (UKB) and All of Us Research Program (AoU), now make STR association studies possible at population scale.

Here, we performed a screen for TREs associated with neurodegenerative and related diseases across two independent biobank cohorts totaling over 320,000 individuals from the UKB and AoU. By combining neurodegenerative diagnoses into composite phenotypes, we increased statistical power, detected pleiotropic TRE loci, and identified candidate disease associations that single-disease analyses lack the power to uncover.

## Methods

### Study populations and ethics statement

Research performed in this study complies with all relevant ethical guidelines, and informed consent for genetic research was obtained from all participants. Collection of UKB data was approved by the Research Ethics Committee of the UKB, obtained under application 82094, and protocols for UKB are overseen by the UKB Ethics Advisory Committee (https://www.ukbiobank.ac.uk/ethics/). Protocols for the AoU cohort are overseen by the All of Us Institutional Review Board (https://allofus.nih.gov/about/who-we-are/institutional-review-board-irb-of-all-of-us-research-program).

We analyzed GS data from 411,916 samples of unrelated European ancestry in UKB and 88,766 samples in AoU across three ancestry groups, European (EUR), African (AFR), and Latino/admixed American (AMR), from the v7 data release. Other ancestry groups were excluded from both cohorts due to insufficient case numbers for case-control analysis. All samples were sequenced using PCR-free Illumina 150 bp paired-end sequencing. Sequencing reads were aligned to the GRCh38 reference genome by both UKB and AoU.^13,14^ Initial QC filters were applied to both cohorts. We removed individuals who had withdrawn consent, failed genome sequencing quality metrics, had predicted sex chromosome aneuploidy, or had second-degree or closer relationships (kinship coefficient > 0.0883); for related pairs, one individual was retained.

European ancestry was determined among UKB participants using PCA of LD-pruned, QC-filtered SNVs (minor allele frequency [MAF] > 0.05, genotyping rate > 99%, Hardy-Weinberg equilibrium [HWE] p > 0.0001, LD pruning) computed with GCTA (v.1.93.2b), using the 1000 Genomes data as the reference panel. A random forest classifier assigned individuals to one of five ancestry groups. Those with assignment probability < 0.5 were excluded. Within-group PCA was then performed to identify and remove ancestry outliers.^15,16^ Individuals with mismatches between self-reported and predicted ancestry, multiple self-reported ancestries across visits, or low assignment probability (p < 0.5) were also excluded. Genetic ancestry was determined for AoU participants (v7) using array-based genotype data by the All of Us Research Program.^14^ Within each ancestry group, centroid distances were computed across 10 PCs as the sum of squared standardized deviations from the group mean, and ancestry outliers were identified by plotting the distribution of centroid distances and removing individuals at the high tail.^15^ Centroid distance thresholds were set to 25 for AFR, AMR, and EUR.

### Short tandem repeat genotyping in UK Biobank and All of Us

We genotyped 22,376 STR loci with ExpansionHunter v5.0.0.^12^ Loci were drawn from a previously described catalog^17^ enriched for polymorphic or expansion-prone TRs, filtered to retain genic and promoter (exonic, intronic, UTR, splicing, or upstream), autosomal loci with motif length ≥ 3 bp and a reference repeat span < 150 bp. Briefly, the catalog was constructed from multiple sources:

1. **Polymorphic TRs** identified by genotyping 2,504 individuals from the 1000 Genomes Project (high-coverage PCR-free Illumina GS) with ExpansionHunter v5.0.0 (motifs 2-20bp) and hipSTR v0.7 (motifs 2-6 bp),^18^ retaining loci with ≥ 20 distinct alleles or ≥ 5 distinct alleles overlapping regulatory or genic regions (ENCODE cCREs, GeneHancer elements, or ANNOVAR-defined exonic, UTR, upstream, downstream or splicing regions).
2. **GC-rich TRs** with motif size 2-10 bp composed entirely of C and G bases, based on Tandem Repeats Finder annotations of GRCh38.^19^
3. **TRs with evidence of expansion**, identified using a combination of ExpansionHunter Denovo v0.9.0,^20^ STRetch v0.4.0,^21^ and hipSTR v0.7 across multiple cohorts including TOPMed, 1000 Genomes Project, Gabriella Miller’s Kids First, Accelerating Medicines Partnership Parkinson’s Disease Initiative, and Pediatric Cardiac Genomics Consortium. Loci were retained if at least one individual showed an outlier expansion.

The final catalog comprised 22,376 STR loci, which were genotyped using CRAM files generated by UKB and AoU (aligned to GRCh38) with ExpansionHunter v5.0.0,^12^ run on the UKB DNAnexus Research Analysis Platform and the All of Us Researcher Workbench, respectively. From the diploid genotypes output by ExpansionHunter, we used the allele size of the longer of the two alleles per sample at each locus in all downstream analyses. This “long-allele” approach was selected to capture a dominant genetic model, which characterizes the vast majority of known pathogenic TREs in neurodegenerative diseases (such as *C9orf72* and *HTT*). Genotypes were quality-filtered to retain autosomal regions with a genotyping rate ≥ 99%, ≥ 2 unique alleles, and a standard deviation > 0. Samples with a genotyping rate < 99%, outlier PCA values, or outlier insert size values were removed. PCA of long-allele genotypes revealed batch effects driven by the sequencing center in UKB and by both the sequencing center and ancestry in AoU.

### Tandem repeat expansion calling

Given the batch effects observed in PCA, we called putative STR expansions independently within subcohorts defined by sequencing center and, in AoU, additionally by ancestry (Table S1). In UKB, samples were divided into two subcohorts by sequencing center: deCODE and the Sanger Center (SC). In AoU, samples were stratified into 12 subcohorts defined by the combination of genetically inferred ancestry (EUR, AFR, or AMR, as described above) and four sequencing batches across three sequencing centers. AoU sequencing batches were defined based on the three contributing sequencing centers (Baylor College of Medicine, University of Washington, and the Broad Institute), with Broad Institute samples further subdivided into two batches based on biosample collection date (before or after September 2019), as we observed a pronounced shift in insert size distribution between these two periods upon inspection of insert size distributions.

Using the long allele genotypes defined above, we calculated per-locus population statistics for each subcohort, including a z-score-based outlier test. We identified expansions using seven threshold levels, increasing in stringency, based on long allele size, the percentile of long allele size in the population, the FDR-adjusted p-value (q-value) from outlier analysis, and the difference from the population median (Table S2). Testing multiple thresholds allows detection of expansions across a range of sizes, since the optimal cutoff varies by locus depending on the repeat unit and its pathogenic range. For each threshold, individuals carrying a long allele fitting the criteria were defined as “expanded,” and all other alleles were “not expanded”, creating a binary genotype at each threshold. To reduce false-positive TRE calls, we applied a random forest classifier to predict whether each expansion call is a true or false positive.^22^ All genotypes identified as false positives by the classifier were removed from further analysis.

### Phenotype definition

We constructed two composite neurodegenerative phenotypes for each cohort. The composite phenotypes are binary, with cases defined as participants with at least one of the included neurodegenerative phenotypes (Tables S3 and S4). The first composite phenotype includes all neurodegenerative and related traits (ND_All_). The second excludes Parkinson’s disease and Alzheimer’s disease (ND_Subset_), since these two diagnoses contribute a large proportion of cases in ND_All_ and could mask signals from other neurodegenerative conditions. Both phenotype definitions and their exclusion lists were manually curated.

In UKB, phenotypes were derived from *International Classification of Diseases*, 10th revision (ICD-10) codes across three data fields: hospital inpatient diagnoses (field 41270), underlying cause of death (field 40001), and self-reported non-cancer illness codes (field 20002), accessed through UKB application no. 82094.^16,17^ Included diagnoses encompassed a broad range of neurodegenerative phenotypes (Table S3). Individuals with secondary or drug-induced Parkinsonism (e.g., ICD-10 codes G21, G22) and vascular dementia were excluded from the case set to ensure the phenotype captured primary neurodegenerative disease (Table S5).

In AoU, phenotypes were derived from OMOP concept codes (v5.3.1) from the conditions domain and from self-reported survey responses, which were converted into binary variables indicating the presence or absence of each response option. We identified and manually curated a set of neurodegenerative phenotypes spanning hereditary, degenerative, and motor neuron diseases from the available binary phenotypes (Table S4). Individuals with secondary parkinsonism, vascular parkinsonism, or vascular dementia were excluded from the case set (Table S5).

Controls were defined as neurologically and psychiatrically healthy older individuals aged ≥ 60 years in UKB and ≥ 50 years in AoU, with no recorded diagnoses of relevant neurological or psychiatric conditions and no use of relevant medications. A younger age cutoff was used in AoU to accommodate the younger demographic distribution of the AoU cohort while maintaining sufficient numbers of controls. In both cohorts, individuals were excluded from the control set if they had recorded diagnoses of neurological or psychiatric conditions related to neurodegeneration, were prescribed relevant medications, or reported a personal or family history of neurodegenerative disease via survey, to minimize the inclusion of prodromal cases or individuals with elevated genetic risk (Tables S6 and S7). Cases and controls are mutually exclusive sets of samples. Individuals that met case criteria were not eligible for inclusion in the control set regardless of age.

After phenotype definition and filtering, the UKB cohort comprised 10,770 cases in ND_All_, 3,759 cases in ND_Subset_, and 265,641 controls. The AoU cohort comprised 4,291 cases in ND_All_, 2,803 cases in ND_Subset_, and 40,079 controls (Table S1).

### Tandem repeat expansion association analysis

To identify TREs associated with the two composite neurodegenerative phenotypes, we used the set of filtered binary genotype calls from the seven TRE thresholds in an association analysis. Of the 22,376 regions genotyped via ExpansionHunter, 6,539 passing strict downstream filter metrics (minor allele count [MAC] ≥ 2 expanded individuals, ≥ 1 expanded case, non-intergenic, and motif length ≥ 3 bp) were advanced to the final association analysis.

Association analysis was performed using REGENIE v3.4.1^23^ in two steps. Step 1 fit the whole-genome regression model to capture polygenic effects and population structure, and Step 2 performed the per-locus association testing. Logistic regression was used for binary outcomes with Firth bias correction due to the unbalanced case-control phenotype, using REGENIE’s approximate Firth correction (--firth --approx) to reduce computational burden while retaining the bias-correcting properties of the exact Firth approach. The Firth correction reduces bias in parameter estimates when rare events or separation in the data are present. Association analyses were run separately in UKB and AoU. Within each cohort, we combined the subcohorts from the expansion calling into a single association test, incorporating the sequencing center as a covariate to account for batch differences. Covariates differed slightly between cohorts, reflecting differences in study design; for UKB, the model was:

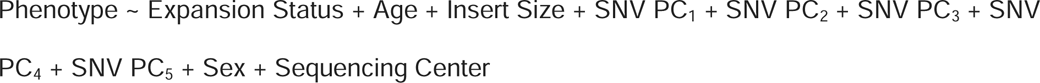

and for AoU:

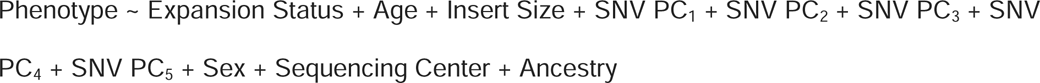

where Phenotype and Expansion Status are binary (0/1) variables, SNV PC_1-5_ are the first five principal components derived from SNV genotype data, and ancestry was included in AoU to account for the multi-ancestry study design. Each combination of expansion threshold (n = 7) and phenotype (n = 2) was analyzed separately, with a per-batch Step 2 size of 1,000 in UKB and 100 in AoU and a minimum MAC of 2.

### Meta-analysis of AoU and UKB

We performed a sample-size-weighted fixed-effects meta-analysis of REGENIE results from UKB and AoU using METAL v2018-08-28.^24^ For each combination of STR locus, phenotype, and expansion threshold, summary statistics from each cohort were combined using the sample-size-weighted meta-analysis scheme in METAL, generating a pooled Z-statistic and meta-analysis p-value for each locus-phenotype-threshold combination. Results were analyzed separately for each of the two phenotype definitions (ND_All_ and ND_Subset_) and the seven thresholds (Table S2). Because tests at different thresholds are highly correlated due to many shared variants and samples, applying a Bonferroni or FDR correction across the thresholds would be overly conservative. FDR correction using the Benjamini-Hochberg method was therefore performed separately for each of the 14 total tests (7 thresholds and 2 phenotypes). For each phenotype, we selected the threshold with the lowest q-value for each locus. We added functional annotations to each region with ANNOVAR.^25^ Significant loci known to cause neurodegenerative diseases were annotated as known pathogenic TRE disorders.

### Validation of expanded genotypes

Candidate loci with q < 0.1 were further evaluated to validate the underlying genotypes. We selected a few expanded samples per locus and generated REViewer^26^ haplotype-resolved visualizations of read alignments to confirm the size of expansion and read support for each genotype call. Loci with insufficient coverage or large mismatches were removed.

To validate ExpansionHunter genotypes at significant loci, we used long-read GS data from the AoU project generated with Pacific Biosciences (PacBio) HiFi technology. We genotyped the same STR loci in the long-read data using TRGT (v1.2),^27^ which uses long-read data to produce more accurate STR genotypes than short-read methods, using the same STR definitions as in ExpansionHunter. TRGT genotypes were quality-filtered by removing those supported by only a single spanning read or with median purity scores from TRGT < 0.75. Primary concordance analyses were performed using TRGT genotypes available at the time of analysis (n = 1,589 individuals). For each individual, we compared the long allele size from ExpansionHunter with that from TRGT at each significant locus and confirmed concordance between the two platforms using Spearman’s correlation. We removed regions with a correlation coefficient of < 0.6. To evaluate the performance of the random forest classifier in removing false positives, we compared outliers identified in EH genotypes (based on short-read Illumina GS) to those identified in TRGT (based on PacBio long-read GS) at the 98^th^ percentile threshold, assuming TRGT genotypes as a gold standard truth set, and calculated precision (i.e. the fraction of EH-called outliers at the 98^th^ percentile that were confirmed as outliers in TRGT) before and after applying the classifier.

The AoU v9 data release subsequently made TRGT genotypes available for an expanded set of 12,223 long-read samples. We calculated long allele size percentiles and mean purity scores of TREs across this broader dataset. For the 3,600 samples with both EH and TRGT genotypes available, we additionally confirmed concordance of outliers at the 98^th^ percentile before and after the random forest classifier at each significant and suggestive locus. For significant candidate loci, we systematically screened all significant carriers for co-occurring expansions at established neurodegenerative loci. Where an individual was found to carry both a pathogenic expansion and a candidate expansion in an additional locus, we assessed whether the statistical signal at the locus could be attributed to the co-occurring expansion.

### Per allele size association

To better assess the pathogenic threshold, we conducted additional testing on any loci that were nominally significant (p<0.05) in the meta-analysis. Because known pathogenic TREs vary widely in their pathogenic size thresholds, the seven predefined expansion thresholds do not always capture the full picture of where a true association begins; this analysis was therefore performed to refine the putative pathogenic threshold at each locus. For each locus, we defined a series of binary thresholds by iterating over every observed allele size above the 99^th^ percentile, classifying individuals with alleles at or above each threshold as expanded and those below as unexpanded. For each of these thresholds, we re-ran the association analysis using Firth logistic regression, the same model and covariates as before, but implemented it with the brglm package in R. We then selected the allele size threshold that yielded the lowest p-value. FDR correction was applied to p-values in each phenotype tested separately.

### Fine-mapping and conditional analysis

To evaluate whether associations were attributable to TREs or to co-inherited single-nucleotide variants (SNVs) that might occur in linkage disequilibrium with the TRE, we performed fine-mapping and conditional analyses separately in each cohort. For each TR-phenotype pair with q < 0.1, we extracted SNV genotypes within a ±250 kb window of the STR locus. SNV data were extracted from whole-genome sequencing data generated by both cohorts. In both UKB and AoU, SNV genotypes were called from PCR-free Illumina 150 bp paired-end GS data aligned to GRCh38 using the Illumina DRAGEN pipeline, with variant calling performed centrally by each respective program.^13,14^ Using PLINK2,^28^ we filtered SNVs to retain those with MAF ≥ 0.01, genotyping rate ≥ 0.95, HWE p-value > 1×10-300, and MAC ≥ 100 in the study population. For AoU, SNV data from EUR, AFR, and AMR samples were merged with PLINK^29^/PLINK2, using allele flipping as needed to resolve strand differences. Linkage disequilibrium (LD) pruning was applied using PLINK2 –indep-pairwise with a window size of 1000 SNVs, step size of 100 SNVs, and LD threshold r² = 0.9 to use in global null-model fitting in Step 1 of REGENIE.

We performed a two-step association analysis with REGENIE, using the same model as the STR analysis in both UKB and AoU, with the extracted SNV data. In Step 1, a global null model was fit using all LD-pruned SNVs genome-wide within each cohort separately (UKB and AoU), consistent with the approach used in the main association analysis. In Step 2, all QC-filtered SNVs in the ±250 kb window of each STR locus were tested for association with the phenotype using the Step 1 output. The top 100 SNVs (ranked by p-value) per region were retained for subsequent fine-mapping and conditional analyses.

Statistical fine-mapping was performed with CAVIAR v2019-04-19.^30^ We extracted the Z-scores of the top 100 SNVs based on the p-value from the SNV association analysis. These SNVs were LD pruned with r² = 0.99 to remove variants in perfect LD. A pairwise LD matrix was generated from the resulting SNVs with PLINK2. The Z-scores and pairwise LD matrix were used as input for CAVIAR. CAVIAR was run with a maximum of 3 causal variants per locus (-c 3), a causal prior probability of 0.01 (-g 0.01), and a 95% credible set threshold (-r 0.95).

Two complementary conditional analyses were performed with REGENIE Step 2 on the top 100 SNVs from the unconditioned SNV association analysis, using the same models and covariates, run separately in UKB and AoU. Results were interpreted per cohort rather than combined via meta-analysis. We used the same global null model fit as the LD-pruned SNVs output from Step 1 above. The two conditional analyses performed in each cohort are:

1. Conditioning on the lead SNV: The lead SNV was defined as the SNV with the lowest p-value from the unconditioned association analysis. The REGENIE test was run separately for each STR locus, including the STR and the other 99 top SNVs in the ±250 kb window, conditioning on the top SNV. This tested whether the TRE association persisted after accounting for the most strongly associated nearby SNV.
2. Conditioning on the TR: The REGENIE test was run separately for each ±250 kb window for the top 100 SNVs, conditioning on the STR variant itself. This was done to test whether the SNV association signal persisted after accounting for the TR.

### cis-eQTL analysis of tandem repeat expansions

To explore potential regulatory mechanisms underlying disease-associated TREs, we tested whether these TRs act as cis expression quantitative trait loci (eQTLs) across three independent cohorts: Genotype-Tissue Expression (GTEx) v8, the Multi-Ethnic Study of Atherosclerosis (MESA), and the Accelerating Medicines Partnership Parkinson’s Disease Initiative (AMP-PD).

In each cohort, ExpansionHunter was used to genotype tandem repeat lengths. Expanded alleles were filtered using the random forest classifier to remove false positives, as described above. The long allele length was used as a continuous predictor. In all three cohorts, expression values were normalized using a rank-based inverse-normal transform (INT) applied per gene across samples, and then residualized on covariates via linear regression. In GTEx, we used their pre-processed *normalized_expression.bed* files, which were then residualized on covariates, including sex, median insert size, sequencer, genotyping principal components, and PEER factors. In MESA, raw transcripts per million (TPM) expression values were INT-transformed within the analysis and then residualized on sex, median insert size, age, analyte isolation batch, and predicted ancestry. In AMP-PD, raw normalized counts were likewise INT-transformed within the analysis, then residualized on sex, age, cohort (study of origin), median insert size, and five genotyping principal components. The resulting residuals served as the expression phenotype in the eQTL model, in identical form across all three cohorts. For each STR locus, all genes that fell within ±100 kb of the STR were tested. The eQTL model used ordinary least squares linear regression of residualized gene expression on the STR long allele length as a continuous predictor, with a minimum of five distinct allele lengths and ten samples per locus–gene–tissue combination required for model fitting.

Across the STR loci tested, a total of 92 unique genes were tested in GTEx (49 tissues; n = 71–683 per tissue; median 70 genes per tissue; 3,401 locus–gene–tissue tests), 94 unique genes in MESA (T-cells n = 333, PBMCs n = 994, monocytes n = 327; 227 locus–gene–tissue tests), and 126 unique genes in AMP-PD (n = 2,195; 126 locus–gene tests). In GTEx, for each locus-gene pair, we computed per-tissue q-values using the Benjamini-Hochberg procedure and defined significance at q < 0.1. We additionally performed cross-tissue random-effects meta-analysis using *metafor::rma* separately for brain tissues (13 tissues), non-brain tissues (36 tissues), and all tissues combined (49 tissues). In MESA, per-compartment q-values were computed as above (q < 0.1 threshold). In AMP-PD, q-values were computed across all results.

## Results

### Association analysis identifies known and candidate tandem repeat expansions associated with neurodegenerative disease

To identify TREs associated with neurodegenerative disease, we performed an association study across 6,539 autosomal STR loci and meta-analyzed results from the UKB and AoU cohorts. Following STR genotyping on GS samples in AoU and UKB, expansions were called at seven different thresholds within subcohorts to account for batch effects of sequencing center and ancestry (Table S2). A random forest classifier was applied to remove false-positive expansions. When comparing EH outliers to TRGT outliers at the 98^th^ percentile, median precision of expansions across loci in genic and promoter regions increased from 0.89 in raw genotypes to 1 after applying the classifier (Figure S1). We tested two composite neurodegenerative phenotypes: all neurodegenerative diseases (ND_All_; 10,770 cases and 265,641 controls in UKB; 4,291 cases and 40,079 controls in AoU; Table S1) and neurodegenerative diseases excluding Parkinson’s disease and Alzheimer’s disease (ND_Subset_; 3,759 cases and 265,641 controls in UKB; 2,803 cases and 40,079 controls in AoU; Table S1). Case and control phenotype definitions and exclusion criteria are detailed in Tables S3-S7.

Across both phenotype analyses, meta-analysis identified multiple loci reaching q < 0.1, including both established pathogenic TRE loci and candidate associations (Figures 1 and 2, Tables S8 and S9). In the ND_All_ tests, 18 loci reached q < 0.1 and survived QC filtering (Figure 1, Table 1). Of these, nine reached q < 0.05. Seven of these are well-established pathogenic neurodegenerative TREs. The three most significant associations were at *C9orf72* (GGGGCC, intronic; q = 1.3×10^-81^), *DMPK* (CAG, splicing; q = 1.1×10^-58^), and *HTT* (CAG, exonic; q = 2.4×10^-9^), which validates the pipeline’s ability to recover TREs associated with neurodegenerative phenotypes. Allele size distributions for ND_All_ known TREs can be found in Figures S2 and S3. Expanded *C9orf72* carriers in both cohorts spanned diagnoses including dementias, ALS, motor neuron disease, ataxia, and unspecified degenerative disorders, consistent with the pleiotropic disease spectrum of *C9orf72* expansions (Table S10). Additional known pathogenic loci reaching q < 0.05 included *ATXN3, CACNA1A, ATXN2*, and *CNBP*. *ATXN2* expanded carriers showed the broadest phenotypic spread, with diagnoses including ALS, cerebellar ataxia, Parkinson’s disease, motor neuron disease, and Alzheimer’s disease in AoU, and hereditary ataxia and motor neuron disease in UKB (Table S10). The two candidate loci at q < 0.05 are *DAPK1* (AATAA, intronic; q = 4.5×10^-3^, Figure 3A, Figure 4A) and *ANK3* (ATTTT, intronic; q = 0.034, Figure 3B, Figure 4B). Additional allele size distributions for ND_All_ candidate TREs can be found in Figures S4 and S5. Ten loci that reached q < 0.1 were excluded in QC where mapped reads did not support the ExpansionHunter genotype upon visual inspection of REViewer plots and/or where the Spearman correlation with TRGT genotypes from long-read data was < 0.6 (Table S11). The *FARP2* locus was also excluded because a co-occurring *ATXN2* expansion better explained the observed phenotype.

**Figure 1.**
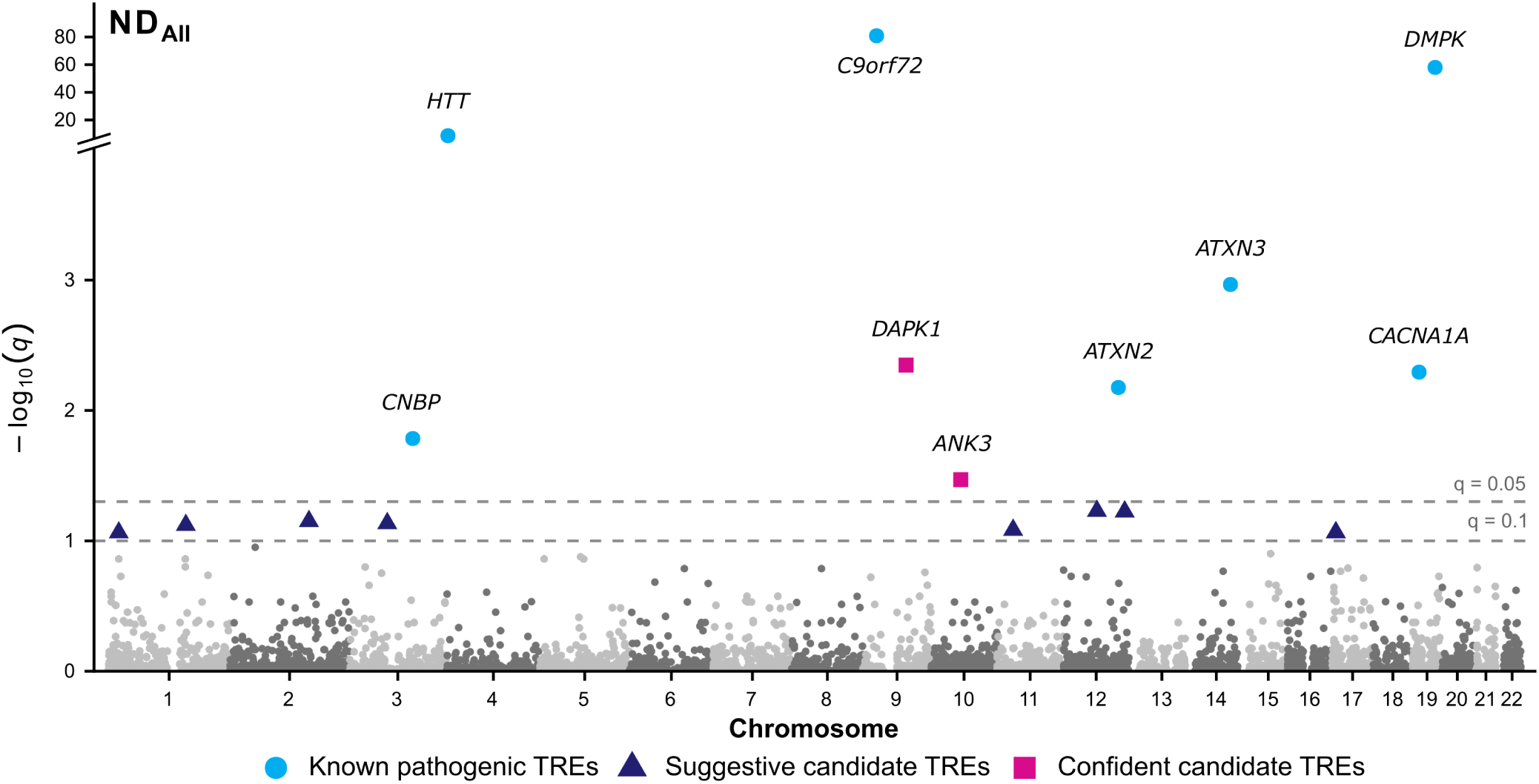
Association analysis results for tandem repeat expansions across all neurodegenerative diseases. Manhattan plot showing −log_10_(q) for association between TREs and the ND_All_ composite phenotype, tested across 6,539 autosomal loci and meta-analyzed across UKB and AoU. Each point represents one STR locus, colored and shaped by classification: known pathogenic TREs (cyan circles), suggestive candidate TREs passing q < 0.1 (navy triangles), and confident candidate TREs passing q < 0.05 (magenta squares). TREs with q < 0.1 that did not pass quality control are not shown. Dashed lines mark q thresholds of 0.05 (upper) and 0.1 (lower). The y-axis is broken to accommodate the highly significant known loci (*C9orf72, DMPK, HTT*).

**Figure 2.**
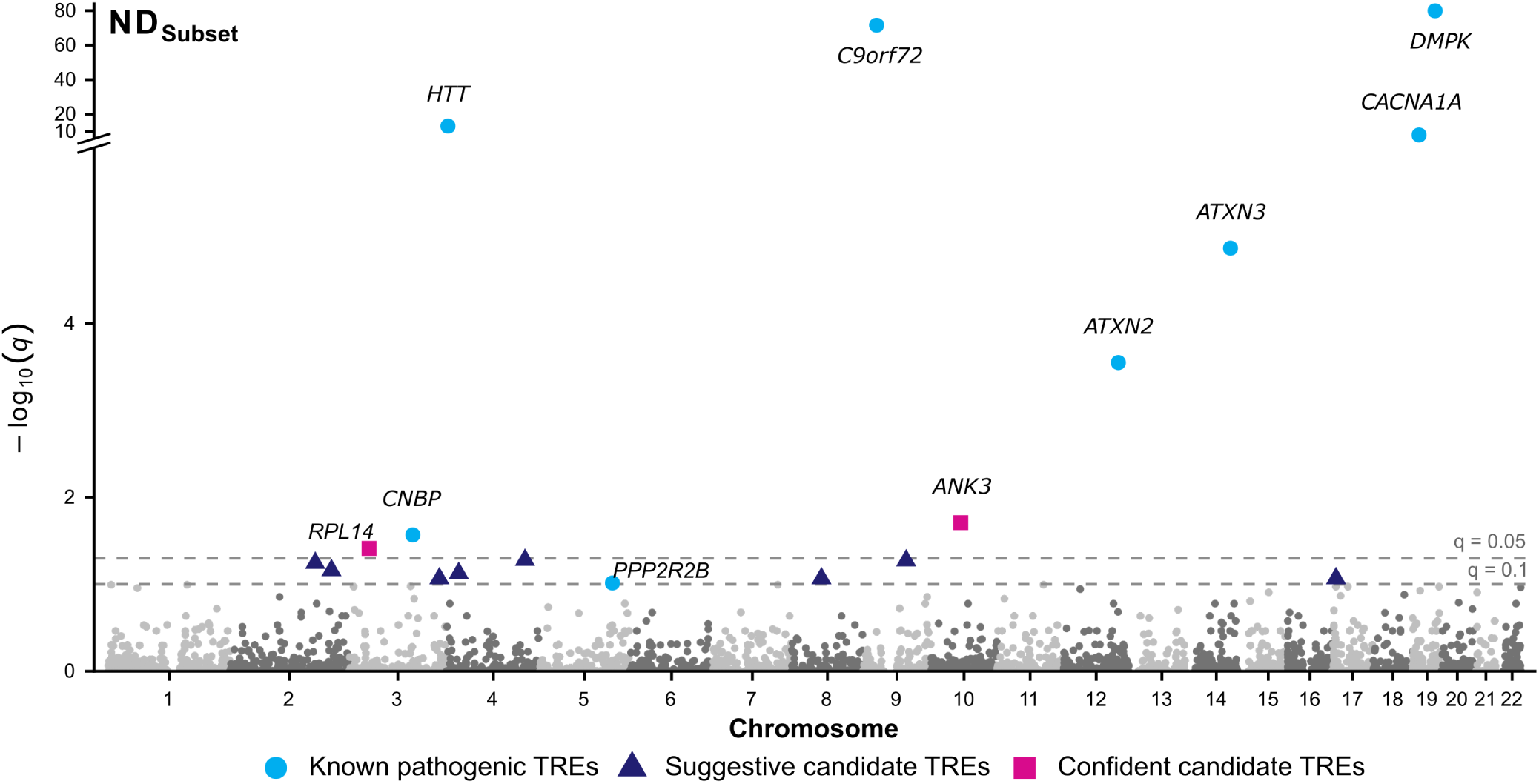
Association analysis results for tandem repeat expansions across neurodegenerative diseases excluding Parkinson’s disease and Alzheimer’s disease. Manhattan plot showing −log_₁₀_(q) for association between TREs and the ND_Subset_ composite phenotype, tested across the same 6,539 autosomal loci and meta-analyzed across UKB and AoU. Colors and shapes follow the same scheme as Figure 1. TREs with q < 0.1 that did not pass quality control are not shown. The y-axis is broken to accommodate the highly significant known loci (*C9orf72, DMPK, HTT, CACNA1A*).

**Table 1.** Meta-analysis results for tandem repeat expansion associations with all neurodegenerative diseases. Loci with q < 0.1 and passing QC are shown for ND_All_, ordered by q-value. Genomic coordinates are in GRCh38.

| Locus (hg38) | Threshold (percentile) | RefUnit | Location | Gene Name | q-value |
| --- | --- | --- | --- | --- | --- |
| chr9:27573528-27573546 | 99.5th | GGGGCC | intronic | *C9orf72*<sup>a</sup> | $1.3 \times 10^{-81}$ |
| chr19:45770204-45770264 | 99.95th | CAG | splicing | *DMPK*<sup>a</sup> | $1.1 \times 10^{-58}$ |
| chr4:3074876-3074933 | 99.9th | CAG | exonic | *HTT*<sup>a</sup> | $2.4 \times 10^{-9}$ |
| chr14:92071009-92071042 | 99.95th | GCT | exonic | *ATXN3*<sup>a</sup> | $1.1 \times 10^{-3}$ |
| chr9:87575213-87575273 | 99.9th | AATAA | intronic | *DAPK1* | $4.5 \times 10^{-3}$ |
| chr19:13207858-13207897 | 99.8th | CTG | exonic | *CACNA1A*<sup>a</sup> | $5.1 \times 10^{-3}$ |
| chr12:111598949-111599018 | 99.95th | GCT | exonic | *ATXN2*<sup>a</sup> | $6.7 \times 10^{-3}$ |
| chr3:129172576-129172656 | 99.8th | CAGG | intronic | *CNBP*<sup>a</sup> | 0.016 |
| chr10:60187128-60187172 | 99.9th | ATTTT | intronic | *ANK3* | 0.034 |
| chr12:68111712-68111780 | 99.5th | AAAAT | ncRNA intronic | *IFNG-AS1* | 0.059 |
| chr12:124402512-124402548 | 99.95th | GCT | exonic | *NCOR2* | 0.060 |
| chr2:160493521-160493593 | 99.95th | TCC | 5'UTR | *RBMS1* | 0.071 |
| chr3:77426678-77426735 | 99.8th | AGGA | intronic | *ROBO2* | 0.073 |
| chr1:157579699-157579749 | 99.5th | ATAC | intronic | *FCRL4* | 0.076 |
| chr11:32990653-32990703 | 95th | AAAT | intronic | *QSER1* | 0.082 |
| chr17:7885025-7885081 | 99.95th | CCG | exonic | *CHD3* | 0.086 |
| chr17:7307649-7307689 | 99.95th | CGG | 5'UTR | *EIF5A* | 0.086 |
| chr1:21527009-21527046 | 99.95th | TTCTT | intronic | *ALPL* | 0.086 |

<sup>a</sup>Known pathogenic TREs

**Figure 3.**
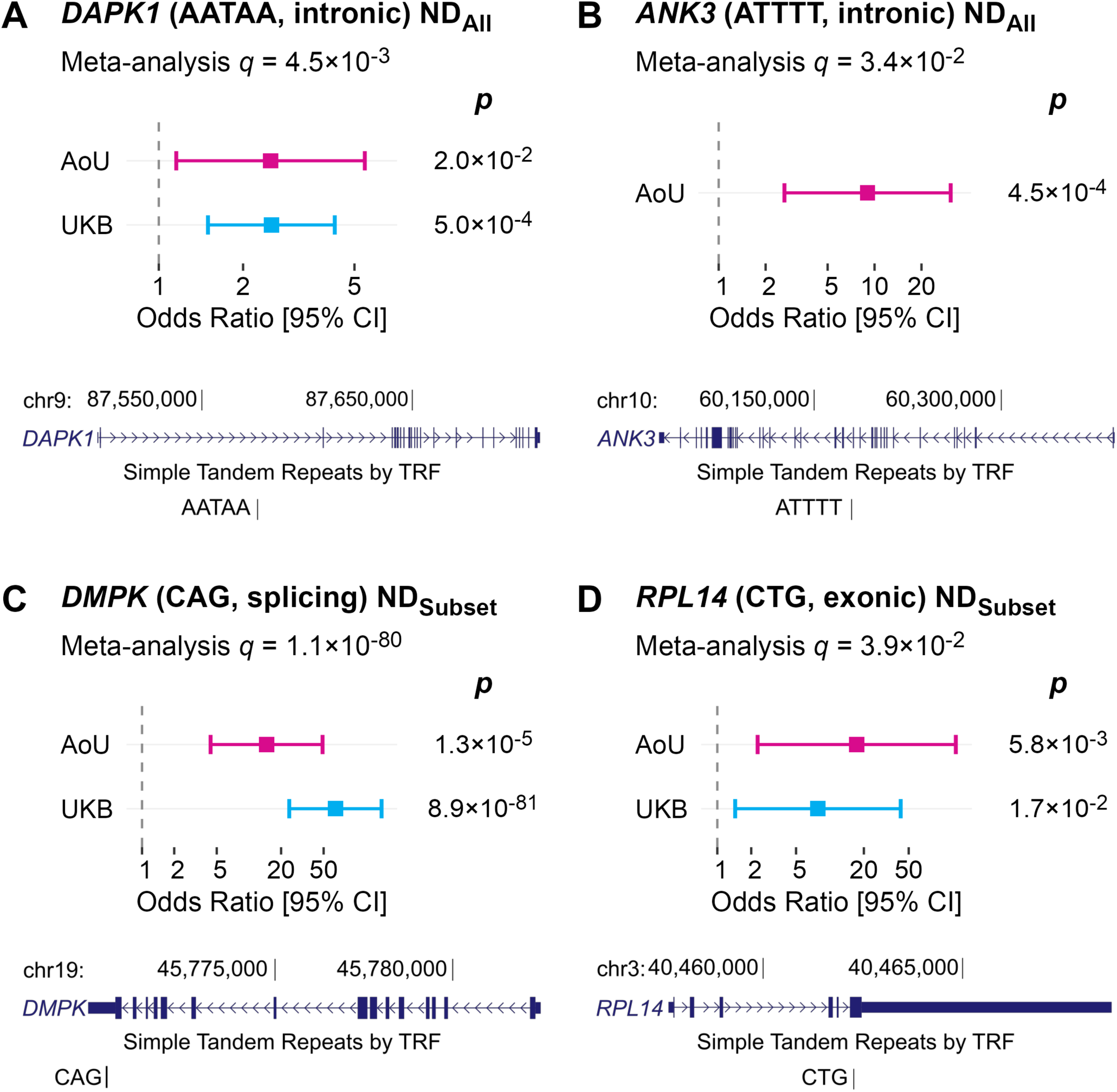
Forest plots and genomic context of tandem repeat expansion associations at four representative loci. Forest plots showing the association between TRE carrier status and the risk of neurodegenerative disease. For each locus, REGENIE was run separately in AoU (magenta) and UKB (cyan), and the two cohort-level results were combined by meta-analysis. (A) *DAPK1* (AATAA repeat, intronic), (B) *ANK3* (ATTTT repeat, intronic), both tested against ND_All_, (C) *DMPK* (CAG repeat, splicing, known pathogenic locus), and (D) *RPL14* (CTG repeat, exonic), both against ND_Subset_. For *ANK3* (B), no expansion carriers were observed in UKB, so only the AoU REGENIE result is shown. In each panel, points show the odds ratio (OR) from each cohort’s REGENIE run, with horizontal lines indicating the 95% confidence interval (CI); the vertical dashed line marks odds ratio (OR) = 1 (no effect). Nominal p-values for each cohort’s REGENIE test are given to the right of each forest panel, and the meta-analysis q-value combining both cohorts is given below each panel’s title. OR and 95% CIs are shown on a log_10_ scale; tick labels reflect the original OR values. Below each forest plot, a UCSC Genome Browser screenshot (hg38, GENCODE V50) shows the genomic context of the repeat locus, with the Simple Tandem Repeats by TRF track indicating the position of the repeat unit.

**Figure 4.**
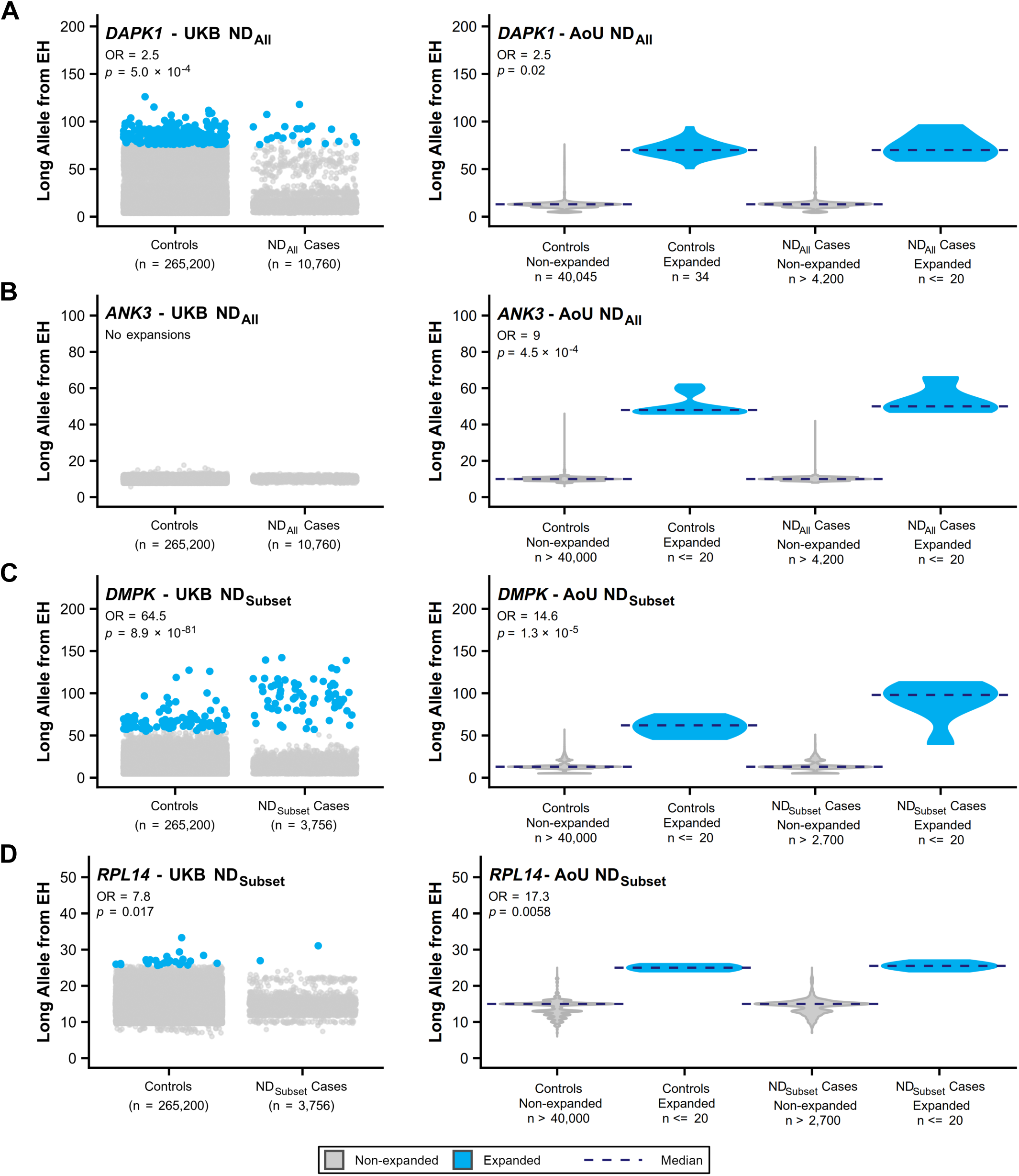
Long allele repeat expansion size at four short tandem repeat loci in UK Biobank and All of Us. (A) *DAPK1*, (B) *ANK3*, (C) *DMPK*, (D) *RPL14*. For each locus, cases are drawn from either the full neurodegenerative-disease cohort (ND_All_) or the neurodegenerative-disease cohort excluding Parkinson’s and Alzheimer’s disease (ND_Subset_), as indicated in the panel title; controls are shared between the two phenotypes. OR and p-value are from the REGENIE association test between case/control status and expansion status in the corresponding cohort (UKB or AoU). Left plots for UKB show individual-level long allele size per participant (grey points, non-expanded alleles; blue points, expanded alleles). Right plots for AoU show the same comparisons in AoU as violin distributions split by case/control and expansion status (grey, non-expanded; blue, expanded), with a dashed navy line marking each group’s median. Because AoU data use policies restrict the release of individual-level data points and exact small-group sizes, AoU results are shown as distributions rather than raw points, and group sizes and near-uniform distributions are obscured as needed, without altering the ORs or p-values. Y-axes are fixed per locus and matched between UKB and AoU for direct comparison.

Notably, all expanded *ANK3* alleles, both cases and controls, were observed exclusively in individuals of AMR and AFR ancestry in AoU, with no expansions detected in EUR samples in either cohort, suggesting that this association may reflect an ancestry-enriched variant not captured in the predominantly European UKB cohort. In this test, *ANK3* expanded carriers in AoU primarily had ataxia, with additional Parkinson’s disease and degenerative disease phenotypes (Table S10). In the ND_All_ analysis, expanded *DAPK1* carriers spanned a broad range of neurodegenerative diagnoses in both cohorts. In AoU, carriers included individuals with ALS, idiopathic progressive polyneuropathy, Parkinson’s disease, cerebellar ataxia, general ataxia, degenerative disorder, degenerative brain disorder, and Alzheimer’s disease. In UKB, carriers were diagnosed (or had cause of death recorded) across an even wider set of codes, including Parkinson’s disease, motor neuron disease, Alzheimer’s disease (early-onset, late-onset, and unspecified), dementia in Pick’s disease, degenerative diseases of the basal ganglia, circumscribed brain atrophy, other degenerative diseases of the nervous system, and unspecified ataxia (Table S10).

In the ND_Subset_ analysis, 18 loci reached q < 0.1 and survived QC filtering (Figure 2, Table 2). Of these, nine reached q < 0.05. Known pathogenic loci again dominated the top associations with *DMPK* (q = 1.2×10^-80^, Figure 3C, Figure 4C), *C9orf72* (q = 2.6×10^-72^), *HTT* (q = 9.1×10^-14^), *CACNA1A* (q = 1.3×10^-8^), *ATXN3* (q = 1.4×10^-5^), and *ATXN2* (q = 2.8×10^-4^) all reaching q < 0.05. Additional size distributions for ND_Subset_ known TREs can be found in Figures S6 and S7. In the ND_Subset_ analysis, carriers of expanded *DMPK* TRs were enriched for diagnoses of muscular dystrophy and myotonic disorders in both cohorts, consistent with its causal role in myotonic dystrophy type 1 (Table S12). *CACNA1A* showed a substantially stronger signal in this phenotype than in ND_All_, consistent with its primary association with spinocerebellar ataxia rather than PD or AD.^31^ Two candidate loci reached q < 0.05: *ANK3* (q = 0.020) and *RPL14* (CTG, exonic; q = 0.039, Figure 3D, Figure 4D), with the latter representing an association not observed in the ND_All_ analysis. *DAPK1* approached but did not reach q < 0.1 (q = 0.053). Additional size distributions for ND_Subset_ candidate TREs can be found in Figures S8 and S9. Expanded *RPL14* carriers in the ND_Subset_ analysis were diagnosed with idiopathic progressive polyneuropathy and degenerative disorder in AoU, and motor neuron disease and unspecified ataxia in UKB (Table S12). 16 loci reached q < 0.1 but were excluded in QC where mapped reads did not support the ExpansionHunter genotype upon visual inspection of REViewer plots and/or where the Spearman correlation with TRGT genotypes from long-read data was < 0.6 (Table S11), and the *FARP2* locus was excluded due to a co-occurring *ATXN2* expansion, as in the ND_All_ analysis.

**Table 2.** Meta-analysis results for tandem repeat expansion associations with neurodegenerative diseases excluding Parkinson’s disease and Alzheimer’s disease. Loci with q < 0.1 and passing QC are shown for ND_Subset_, ordered by q-value. Genomic coordinates are in GRCh38.

| Locus (hg38) | Threshold (percentile) | RefUnit | Location | Gene Name | q-value |
| --- | --- | --- | --- | --- | --- |
| chr19:45770204-45770264 | 99.95th | CAG | splicing | *DMPK*<sup>a</sup> | $1.2 \times 10^{-80}$ |
| chr9:27573528-27573546 | 99.5th | GGGGCC | intronic | *C9orf72*<sup>a</sup> | $2.6 \times 10^{-72}$ |
| chr4:3074876-3074933 | 99.9th | CAG | exonic | *HTT*<sup>a</sup> | $9.1 \times 10^{-14}$ |
| chr19:13207858-13207897 | 99.8th | CTG | exonic | *CACNA1A*<sup>a</sup> | $1.3 \times 10^{-8}$ |
| chr14:92071009-92071042 | 99.95th | GCT | exonic | *ATXN3*<sup>a</sup> | $1.4 \times 10^{-5}$ |
| chr12:111598949-111599018 | 99.8th | GCT | exonic | *ATXN2*<sup>a</sup> | $2.8 \times 10^{-4}$ |
| chr10:60187128-60187172 | 99.95th | ATTTT | intronic | *ANK3* | 0.020 |
| chr3:129172576-129172656 | 99.95th | CAGG | intronic | *CNBP*<sup>a</sup> | 0.027 |
| chr3:40462029-40462059 | 99.9th | CTG | exonic | *RPL14* | 0.039 |
| chr4:159342526-159342616 | 99.95th | TTTTA | intronic | *RAPGEF2* | 0.053 |
| chr9:87575213-87575273 | 99.95th | AATAA | intronic | *DAPK1* | 0.053 |
| chr2:173256480-173256553 | 99.9th | ATAG | splicing | *MAP3K20-AS1* | 0.057 |
| chr2:206249864-206249889 | 99.9th | CTTTT | ncRNA intronic | *GPR1-AS* | 0.069 |
| chr4:24906364-24906418 | 99.9th | ATTTT | intronic | *CCDC149* | 0.074 |
| chr17:7885025-7885081 | 99.95th | CCG | exonic | *CHD3* | 0.086 |
| chr3:183106482-183106510 | 99.95th | TTTC | intronic | *MCCC1* | 0.086 |
| chr8:60678764-60678812 | 99.95th | GCG | 5'UTR | *CHD7* | 0.086 |
| chr5:146878727-146878757 | 99.9th | GCT | 5'UTR | *PPP2R2B*<sup>a</sup> | 0.097 |

<sup>a</sup>Known pathogenic TRES

Nine additional loci for each of the phenotypes reached suggestive significance (q < 0.1, Tables 1 and 2). In the ND_Subset_ analysis, *PPP2R2B* (GCT, 5’UTR; q = 0.097) approached significance, driven by UKB signal (Odds ratio [OR] = 6.50, p = 9.6×10^-4^), with no expanded cases observed in AoU. The GCT repeat we defined at this locus is the same CAG repeat (in reverse complement) previously shown to cause spinocerebellar ataxia type 12 (SCA12).^32^ Expanded UKB carriers were diagnosed with hereditary ataxia, motor neuron disease, and other degenerative diseases of the basal ganglia and nervous system (Table S12), consistent with the known pathogenicity of this TRE. *RAPGEF2* (TTTTA, intronic; q = 0.053) also reached suggestive significance in the ND_Subset_ analysis, driven by UKB (OR = 9.60, p = 6.0×10^-4^), with no signal in AoU. Expanded UKB carriers were diagnosed with dementia in Pick’s disease, hereditary ataxia, circumscribed brain atrophy, and other degenerative diseases of the nervous system (Table S12). This locus overlaps with the repeat region implicated in familial adult myoclonic epilepsy type 7 (FAME7), where an inserted TTTCA pentanucleotide expansion within a background of TTTTA repeats is thought to be the pathogenic motif.^33^ Notably, the cases contributing to this signal had no recorded epilepsy diagnoses, and the phenotypic spectrum differs markedly from the myoclonic epilepsy associated with FAME7, suggesting this may represent a distinct neurodegenerative signal at this locus. Finally, *CHD3* (CCG, exonic; q = 0.086) showed a suggestive association in both phenotype analyses, with the signal restricted to UKB. In the ND_All_ analysis, expanded UKB carriers included individuals with Parkinson’s disease and Alzheimer’s disease diagnoses, as well as hereditary ataxia and other degenerative nervous system diseases. In the ND_Subset_ analysis, carriers were limited to hereditary ataxia and degenerative disease diagnoses (Tables S11 and S12).

Long allele size percentiles and mean purity of the top 20 longest alleles were calculated for the AoU v9 long-read sequencing release (12,223 samples; Table S13). Mean purity was high (≥95%) across most loci, except for *CHD3*, where the 20 longest alleles had a mean purity of 88%. Thus, nearly all observed expansions consisted of uninterrupted reference motif sequence. Among the candidate loci, *DAPK1* showed the widest expansion range, with a 99th percentile of 122 copies and a 99.8th percentile of 198 copies relative to a population median of 12 copies, confirming the presence of large expansions at this locus in long-read data. *RAPGEF2* also showed evidence of large expansions, with a 99.8th percentile of 124 copies compared to a median of 18. Additionally, we assessed the 3,600 samples with both EH genotypes from short-read GS and TRGT genotypes from long-read data (Table S13). Precision of expansion calls at the 98^th^ percentile was high at most loci after classifier-based filtering, with notable improvements at *DAPK1* (0.86 to 1.00), *GPR1-AS* (0.82 to 1.00), and *RAPGEF2* (0.69 to 0.86). At *DAPK1*, EH percentile sizes were smaller than TRGT at the upper tail (99.5^th^ percentile: EH = 52, TRGT = 154 copies), consistent with known underestimation of large expansions by short-read genotyping. *RPL14* showed high concordance across all percentiles and a high true positive rate (Precision = 0.98).

### Per-allele-size analysis refines thresholds at significant loci

To refine the pathogenic size threshold at each associated locus, we performed per-allele-size association testing at all nominally significant loci (METAL p < 0.05; Figures S10- S13, Tables S14 and S15). For *DMPK*, a known pathogenic locus, the per-allele analysis for ND_Subset_ identified thresholds of 57 copies in UKB (OR = 71.2, q = 4.1×10^-132^; Figure S12) and 74 copies in AoU (OR = 50.7, q = 1.2×10^-4^; Figure S13) as the strongest associations, with odds ratios increasing steeply above these thresholds. For *DAPK1*, the sizes that yielded the strongest association were 92 copies in UKB (OR = 5.01, q = 2.1×10^-3^; Figure S11) and 83 copies in AoU (OR = 9.33, q = 0.064; Figure S11) for ND_All_. For *ANK3*, the association was driven by AoU alone, with the strongest association at 47 copies (OR = 9.46, q = 2.9×10^-3^; Figure S11) for ND_All_. For *RPL14*, consistent thresholds were identified as the strongest association in both cohorts at 27 copies in UKB (OR = 13.2, q = 5.2×10^-3^; Figure S13) and 25 copies in AoU (OR = 7.38, q = 0.032; Figure S13) for ND_Subset_. The association reached significance in both cohorts independently.

### Fine-mapping and conditional analyses support TREs as likely variants underlying observed associations

To evaluate whether associations were driven by the TRE itself or by co-inherited SNV variation, we performed CAVIAR fine-mapping and conditional analyses at all loci with q < 0.1 (Tables S16-S21). Among the known pathogenic loci in the ND_All_ phenotype, CAVIAR assigned posterior inclusion probabilities (PIPs) of 1.0 to the TRs at *C9orf72* (Figure 5A) and *HTT* in UKB, and PIPs of 0.99 and 1.0 to *DMPK* in AoU (Figure 5B) and UKB (Figure S14), respectively, confirming the STR as the overwhelmingly likely variant underlying the association at these loci (Table S16). *ATXN3* received a PIP of 0.96 in AoU and *CACNA1A* received a PIP of 0.99 in UKB, further supporting TRE-driven association at these established loci (Figure S14). *ATXN2* showed a high PIP in AoU (0.93; Figure S14) but a markedly lower PIP in UKB (0.03), where a nearby SNV received the top ranking instead. Among candidate loci, *DAPK1* showed CAVIAR support in the ND_All_ analysis in UKB, where the STR ranked as the top variant (PIP = 0.41; Figure 5C). For *ANK3*, CAVIAR results were available in AoU for both phenotype definitions. In the ND_All_ analysis, the STR received the highest PIP among all variants tested (PIP = 0.63; Figure 5D). In the ND_Subset_ analysis, the STR PIP was somewhat lower (PIP = 0.45), though the STR still ranked second among tested variants.

**Figure 5.**
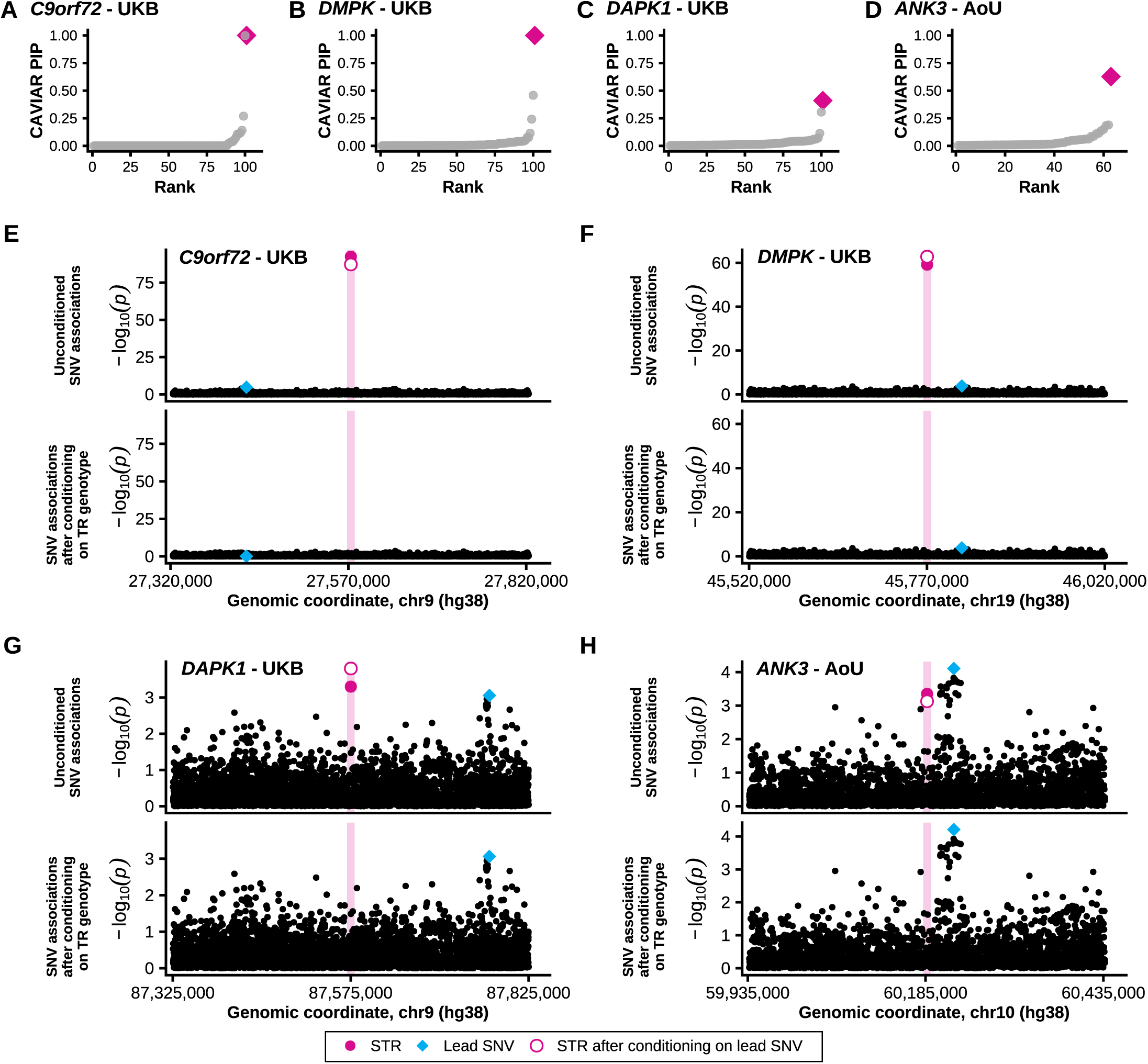
CAVIAR fine-mapping and conditional association evidence at four neurodegenerative disease-associated tandem repeat loci. (A–D) CAVIAR posterior inclusion probability (PIP) for each variant in the fine-mapped credible set at *C9orf72* (UKB), *DMPK* (UKB), *DAPK1* (UKB), and *ANK3* (AoU), respectively. Variants are ranked by ascending PIP (x-axis); grey circles denote SNVs, and the pink diamond denotes the TR. (E–H) Regional association plots for the same four loci (*C9orf72*–UKB, *DMPK*–UKB, *DAPK1*–UKB, *ANK3*– AoU), each shown as two stacked panels. Top: unconditioned associations (−log_10_p) across the region; bottom: SNV associations after conditioning on the STR genotype. In both panels, black points are SNVs; the filled magenta circle marks the TR’s own association, and the open magenta circle marks the TR’s association after conditioning on the locus’s lead SNV; the cyan diamond marks the lead SNV. The shaded pink band indicates the genomic position of the TR. Genomic coordinates are given in GRCh38/hg38. All associations shown (A–H) are from association tests of the NDAll phenotype performed in REGENIE.

Conditional analyses in the ND_All_ analysis provided a clear example of bidirectional confirmation of STR causality at *C9orf72* in UKB. Conditioning on the STR reduced the lead SNV association from highly significant to null (unconditioned p = 1.7×10^-5^; conditioned p = 0.83; Figure 5E, Table S19), while conditioning on the lead SNV left the STR association essentially unchanged (unconditioned p = 3.7×10^-93^; conditioned p = 6.1×10^-88^; Figure 5E; Table S18). This pattern is expected when the STR itself is the causal variant. In *DMPK*, the STR signal was stronger after conditioning on the lead SNV in both UKB (unconditioned p = 6.3×10^-60^; conditioned p = 1.4×10^-63^; Figure 5B) and AoU (unconditioned p = 2.5×10^-4^; conditioned p = 6.5×10^-5^; Figure S16). The association of the lead SNV remained unchanged after conditioning on the STR in both AoU (unconditioned p = 2.2×10^-3^; conditioned p = 2.2×10^-3^; Figure S16) and UKB (unconditioned p = 1.5×10^-4^; conditioned p = 1.4×10^-4^; Figure 5F). In *DAPK1*, the STR association remained significant after conditioning on the lead SNV in both UKB (unconditioned p = 5.01×10^-4^; conditioned p = 1.6×10^-4^; Figure 5G; Table S18) and AoU (unconditioned p = 0.020; conditioned p = 0.018; Table S18). In *ANK3*, the STR association remained essentially unchanged after conditioning on the lead SNV in AoU (unconditioned p = 4.5×10^-4^; conditioned p = 7.9×10^-4^; Figure 5H; Table S18).

For *RPL14* in the ND_Subset_ analysis, CAVIAR PIPs for the STR were low in AoU (0.064), indicating that the current fine-mapping analysis does not fully resolve the variant underlying the association at this locus, consistent with the limited number of expanded carriers at this locus (Table S17).

### *Cis*-eQTL analysis

To evaluate candidate regulatory mechanisms underlying disease-associated TREs, we tested whether disease-associated TRs act as eQTLs for their host and nearby genes. We performed this analysis in 49 tissues in GTEx (Tables S22 and S23), three cell types in MESA (Table S24), and whole blood from AMP-PD (Table S25).

Longer *RPL14* STR alleles were associated with reduced *RPL14* expression in a cross-tissue meta-analysis across all 49 GTEx tissues (β=-0.02 [95% CI: −0.03, −0.02], q=3.1×10^-21^; Figure 6A; Table S23), with 25 of 49 individual tissues reaching FDR significance (Table S22), including tibial nerve (β=-0.02, q=5.0×10^-4^; Figure 6B). The brain-only meta-analysis (k=13 tissues) was directionally consistent but did not reach significance (q=0.15; Table S23). The negative dose-response relationship was replicated in MESA PBMCs, with a larger effect size compared to tissues in GTEx (β=-0.10 [95% CI: −0.13, −0.08], q=5.3×10^-10^; Figure 6C). AMP-PD whole blood showed the same negative direction but did not reach significance (q=0.97; Figure 6A).

**Figure 6.**
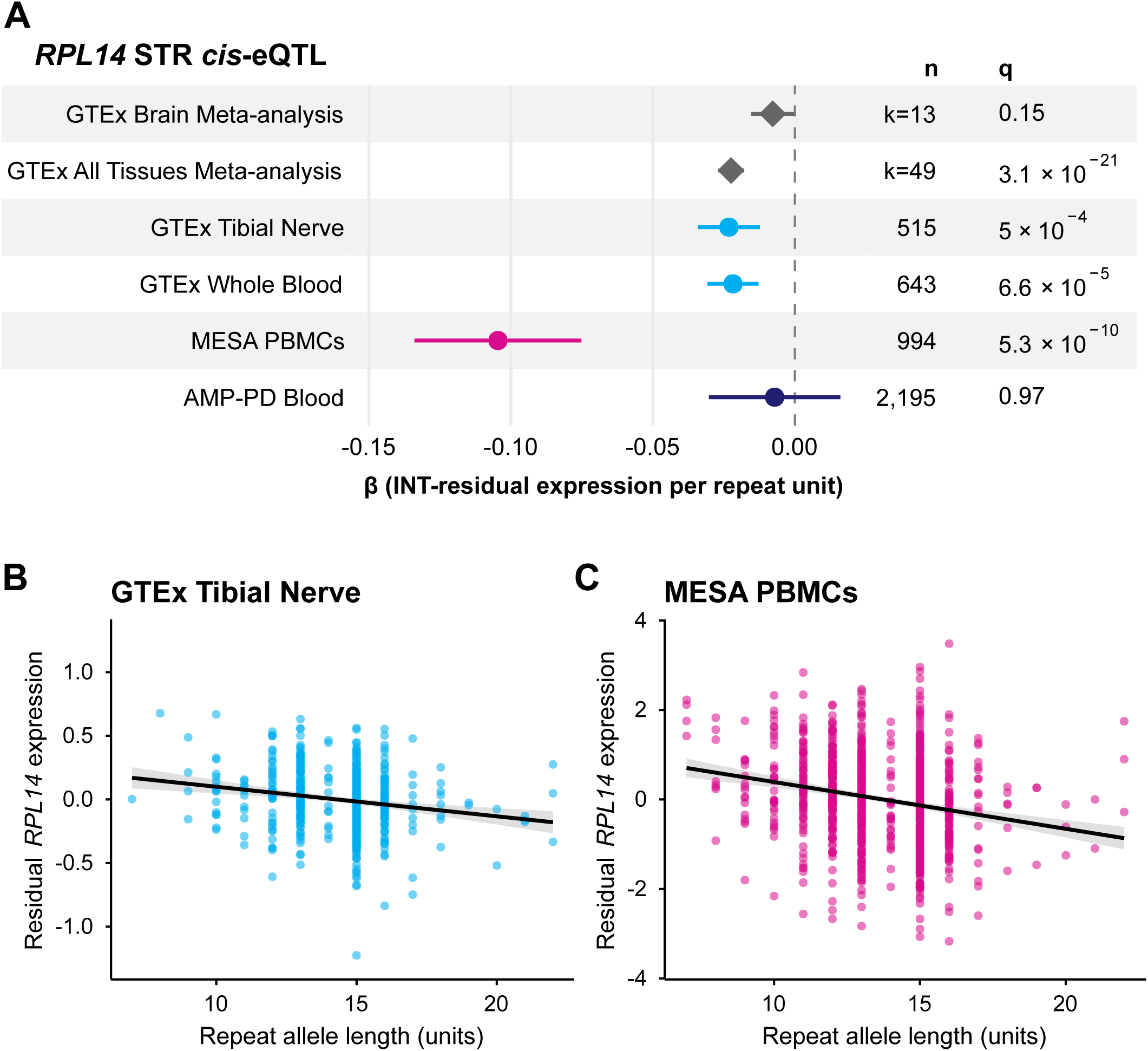
*RPL14* repeat length is associated with *RPL14* expression across tissues and cohorts. (A) Forest plot of the effect (β, repeat units per unit change in INT-residualized expression) of the *RPL14* repeat locus (chr3:40,462,029-40,462,059) on *RPL14* expression, with 95% CI, sample size (n), and q-value, across: a GTEx cross-tissue brain meta-analysis, a GTEx all-tissue meta-analysis, the two GTEx tissues with the strongest signal (Nerve - Tibial and Whole Blood), MESA PBMCs, and AMP-PD blood. For the two meta-analyses, k represents the number of tissues included. Diamonds denote meta-analysis estimates; circles denote single-tissue/cohort estimates. (B–C) Scatter plots of per-sample residual *RPL14* expression (z-score or INT-residual, y-axis) versus repeat allele length (x-axis) in (B) GTEx Tibial Nerve and (C) MESA PBMCs, colored by cohort (GTEx, blue; MESA, pink). Black line, linear fit; grey ribbon, 95% confidence interval of the fit. Sample sizes, effect sizes, and q-values for each of these three tissues/cohorts are given in the corresponding rows of the panel A table.

The *RPL14* STR was also associated with expression of three nearby genes across multiple datasets (Tables S22, S24, S25). *ENTPD3* showed significant associations in 5 GTEx tissues including cerebellum (q=0.022) and cerebellar hemisphere (q=0.048; Figure S17A), though not in MESA or AMP-PD. *ENTPD3-AS1*, the antisense transcript of *ENTPD3*, was negatively associated with the STR in 25 GTEx tissues, MESA PBMCs (q=8.3×10^-9^), and AMP- PD whole blood (q=3.9×10^-26^; Figure S17B). *ZNF619* was negatively associated in 4 GTEx tissues, including cerebellar hemisphere (q=0.008) and MESA PBMCs (q=0.019; Figure S17C). Together, the *RPL14* STR shows a consistent negative effect on expression of its host gene across tissues and cohorts, with additional effects on neighboring genes.

## Discussion

In this study, we performed a tandem repeat expansion association study of combined neurodegenerative diseases, leveraging GS data from 276,411 UKB and 44,370 AoU participants across two composite neurodegenerative phenotypes. By using biobank-scale sample sizes, a curated catalog of 6,539 STR loci, and multi-threshold expansion calling, we identified and validated associations at seven known pathogenic TRE loci and identified additional associations, including at *DAPK1*, *ANK3*, and *RPL14*. The analysis recovered known pathogenic TRE loci, including *C9orf72*, *DMPK*, *HTT*, *ATXN2*, *ATXN3*, *CACNA1A*, and *CNBP*, which are well-established causes of neurodegenerative and related diseases. This confirms that the pipeline recovers genuine biological signals from short-read GS data at scale. At several established loci, the inferred expansion thresholds overlapped with clinically recognized pathogenic ranges, providing additional evidence that the association signals reflect biologically meaningful repeat expansions. For example, the identified thresholds for *HTT* (40-41 repeats), *ATXN2* (32-38 repeats), and *CACNA1A* (21-22 repeats) all occur at or close to their known pathogenic thresholds: ≥40 repeats in *HTT* for full-penetrance Huntington’s disease, ≥33 repeats in *ATXN2* for SCA2 (with intermediate ALS risk at 27-33 repeats), and ≥20 repeats in *CACNA1A* for SCA6.^34–36^

Among the candidate findings, the *DAPK1* intronic AATAA expansion is compelling Death-associated protein kinase 1 (*DAPK1*) is a calcium/calmodulin-regulated serine/threonine kinase with well-established roles in neuronal apoptosis, autophagy, and synaptic plasticity.^37,38^ It has previously been implicated in Alzheimer’s disease, Parkinson’s disease, and Huntington’s disease through both genetic and functional studies.^39^ Dysregulation of *DAPK1* has also been linked to Parkinson’s disease, including evidence that *DAPK1* promotes parkin degradation and increases neuronal vulnerability to neurotoxic insult.^40,41^

The *DAPK1* locus was first implicated in AD through a candidate gene study identifying two intronic SNVs, rs4878104 and rs4877365, in significant association with late-onset AD.^42^ Both SNVs were further associated with allele-specific *DAPK1* expression. Notably, rs4878104 lies 2.8 kb from the AATAA STR identified in the present study, and rs4877365 lies 34 kb from it, placing the expansion in proximity to the previously associated region. The same variant, rs4878104, was subsequently associated with FTD in an Italian case-control study, further implicating *DAPK1* across neurodegenerative diseases.^43^ Our fine-mapping and conditional analysis identified the *DAPK1* expansion as the highest-ranked variant by posterior inclusion probability and by unconditioned significance, with the association remaining significant after conditioning on the lead SNV. This supports a direct role for the repeat itself, consistent with an independent, repeat-mediated contribution at this locus.

The expansion reached FDR significance in the ND_All_ phenotype and suggestive significance in the ND_Subset_ phenotype (excluding AD and PD cases), raising the possibility that *DAPK1* contributes to a broader spectrum of neurodegeneration beyond its established roles in AD and PD. Consistent with this, expansion carriers presented across a broad diagnostic spectrum encompassing ALS, motor neuron disease, cerebellar and general ataxia, idiopathic progressive polyneuropathy, dementia in Pick’s disease, degenerative diseases of the basal ganglia, circumscribed brain atrophy, and unspecified degenerative brain disorders, in addition to AD and PD. This is consistent with the known pleiotropy of *DAPK1* across neurodegenerative diseases and supports the rationale for a composite ND phenotype in this study.

The *ANK3* ATTTT intronic expansion is a particularly notable finding given its ancestry-specific distribution. All expanded alleles in AoU were observed exclusively in individuals of AMR and AFR ancestry, with no expansions detected in EUR samples, potentially explaining the absence of signal in the predominantly European UKB cohort. This underscores the importance of ancestrally diverse cohorts for genetic discovery. The expansion at this locus was significant in both the ND_All_ and ND_Subset_ phenotypes and was the top-ranked variant by CAVIAR PIP in the ND_All_ analysis. While not the top-ranked variant in the regional conditional analysis, the association p-value was not substantially attenuated after conditioning on the lead SNV, suggesting an independent contribution of the repeat.

Expansion carriers in AoU primarily presented with ataxia, with additional Parkinson’s disease and degenerative disease phenotypes. *ANK3* encodes ankyrin-G, a scaffolding protein critical for the organization of the axon initial segment and nodes of Ranvier, where it orchestrates the localization of voltage-gated ion channels and GABAergic presynaptic terminals essential for action potential generation and propagation.^44,45^ *ANK3* has been implicated in bipolar disorder, schizophrenia, and autism spectrum disorder through multiple independent common and rare variant studies, but its role in neurodegeneration has not been established.^44,45^ This locus warrants functional follow-up to clarify whether *ANK3* expansions contribute to neurodegeneration in addition to the gene’s established psychiatric associations.

The exonic CTG repeat in *RPL14* was significant in the ND_Subset_ phenotype, with nominal significance in both cohorts independently and concordant effect directions. This repeat encodes a polyalanine tract, and the observed expansions are modest in size, as expected due to the higher constraint of TRs in exonic regions.^46^ While polyalanine expansions are a well-established cause of multiple TRE-associated diseases, they have previously been associated with congenital disorders and typically impact transcription factors, rather than with neurodegenerative diseases like polyglutamine expansions.^47^ One limitation is that the repeat shows weak support from fine-mapping and conditional analyses. This is likely due to the small number of expansion carriers, which reduces power in LD-based analyses that rely on correlations between the repeat and surrounding variants to identify the causal variant. Even some of the TREs known to be associated with neurodegenerative diseases had weak evidence in those analyses, so it does not rule out the repeat underlying the association.

However, *RPL14* was among the 12 coding CAG loci with elevated Pure Length Variability Index (PLVI, a measure of repeat instability based on longest pure segment length) that replicated across discovery and validation cohorts in a population-scale long-read sequencing study of All of Us participants, flagging it as a candidate repeat expansion disorder, though the authors note that the PLVI pathogenicity signature was not validated for polyalanine loci.^48^ The STRs in *DAPK1* and *ANK3* were in the top 1% and 5% of PLVI scores, respectively.

eQTL analyses further supported this association. Longer alleles of the STR in *RPL14* were negatively correlated with *RPL14* expression in a variety of tissues, including in tibial nerve in GTEx and PBMCs in MESA. The *RPL14* STR is cataloged as a high-confidence fine-mapped expression STR (FM-eSTR) by Fotsing et al.,^49^ who reported a negative association between average repeat length and *RPL14* expression in tibial nerve tissue (β=−0.36, p=3.8×10^-9^) with a CAVIAR posterior probability >0.999 that the TR, rather than a linked SNV, is the likely causal variant driving the expression association.^49^ This was one of only 11 FM-eSTRs identified in coding exons genome-wide. That analysis used HipSTR for repeat genotyping, whereas a subsequent analysis from our group used ExpansionHunter genotypes with a catalog similar to the one used in the present study.^17^ It replicated the negative association between *RPL14* repeat length and expression across GTEx tissues. It also identified associations between *RPL14* repeat length and expression of nearby genes including *ENTPD3*, *ENTPD3-AS1*, and *ZNF619*.

In the present study, we extended this analysis in several ways. Using long allele length rather than average repeat length, we also observed a consistent negative association with *RPL14* expression in GTEx tibial nerve, as well as an additional similar effect in MESA PBMC expression. Meta-analysis across all GTEx tissues and across non-brain GTEx tissues confirmed a highly reproducible signal. No individual brain tissue reached significance, which may reflect limited power given smaller per-tissue sample sizes rather than a true absence of effect. Despite this, the tibial nerve association may be relevant, as *RPL14* expansion carriers included the underlying phenotypes of peripheral neuropathy and motor neuron disease, both of which involve peripheral nerve dysfunction. Additionally, *RPL14* long allele length was associated with changes in the expression of nearby genes such as *ENTPD3*, *ENTPD3*-*AS1*, and *ZNF619* in GTEx, including in some brain tissues. We additionally identified associations between the STR and expression of these genes in the MESA and AMP-PD cohorts. Together, these findings from independent cohorts, genotyping approaches, and tissue contexts provide convergent evidence for a potential functional role of the *RPL14* repeat.

Among the suggestive findings, *RAPGEF2* is of particular interest. The locus we identified overlaps with the repeat region implicated in familial adult myoclonic epilepsy type 7 (FAME7), where an inserted TTTCA pentanucleotide expansion within a background of TTTTA repeats is the likely pathogenic motif.^33^ As ExpansionHunter genotypes the reference TTTTA motif rather than the non-reference TTTCA insertion, it is unclear whether the signal observed here reflects the same pathogenic mechanism. Nevertheless, expanded carriers in this study presented with neurodegenerative phenotypes including dementia, hereditary ataxia, and circumscribed brain atrophy, with no recorded epilepsy diagnoses. This raises the possibility that variation at this locus may contribute to a broader spectrum of neurological disease beyond epilepsy, though replication in larger cohorts will be needed to establish this. Several other loci reaching suggestive significance could be candidates for follow-up, including *CHD3* and *NCOR2*, both of which are exonic repeats, and *CHD7* and *EIF5A*, which harbor GCG and CGG 5’UTR expansions, respectively, a repeat class and location with established disease relevance across multiple neurological diseases.

One of the biggest limitations with this study is the accuracy of the STR genotypes, which is a challenge when using short-read sequencing data. We address this by applying rigorous QC to the ExpansionHunter genotypes and expansion calls, including using our random forest classifier tool to identify false positives, assessing concordance with long-read sequencing genotypes in AoU, and assessing REViewer alignment plots. Concordance between EH and TRGT genotypes in up to 3,600 samples with both short-read and long-read data showed that true positive rates were high at most loci after classifier-based filtering, with notable improvements at *DAPK1* (0.86 to 1.00), *GPR1*-*AS* (0.82 to 1.00), and *RAPGEF2* (0.69 to 0.86). *RPL14* showed high concordance across all percentiles and a high true positive rate (0.98). Additional support for the validity of our *DAPK1* genotypes comes from the newest AoU data release of TRGT genotypes for 12,000 long-read sequencing samples and the Human Pangenome Reference Consortium (HPRC) long-read data. In AoU, the 99.8^th^ percentile long allele size is 198 copies compared to a median long allele length of 12 copies. HPRC has alleles up to 187 copies observed and a 99th percentile of 115 copies, compared to a population median of 10 copies.^50^ These observations confirm the presence of unusually long STR alleles at this locus. In the approximately 3,600 individuals with both EH and TRGT genotypes, *DAPK1* percentiles were concordant at the population level (median: EH = 12, TRGT = 12) but diverged sharply at the upper tail (99.5^th^ percentile: EH = 52, TRGT = 154 copies), directly demonstrating that EH identifies expanded carriers but underestimates their true allele size. This is consistent with the known tendency of ExpansionHunter to correctly detect large expansions while underestimating their precise size when the repeat exceeds the fragment length, as has been documented at other loci when compared to PCR-based or long-read measurements.^51^ Our expansion calls for large expansion sizes could therefore be interpreted as conservative estimates of true allele length.

The power to detect associations is also limited by the number of expansion carriers at a given locus, meaning some genuine disease-associated expansions may remain undetected even at biobank scale. The use of composite neurodegenerative phenotypes increases power by increasing case counts and enables the study of pleiotropy, but it can also dilute disease-specific association signals and obscure which specific phenotypes underlie the significant associations. It is also biased to phenotypes with larger sample sizes. Additionally, because UKB is primarily composed of individuals of European descent, this limits its generalizability and its power to detect ancestry-specific associations. However, the inclusion of AMR and AFR participants in AoU partially addresses this, as seen in the *ANK3* association driven by those participants and not the European participants in either AoU or UKB.

Overall, this study highlights several complementary approaches that together strengthen the evidence for TRE contributions to neurodegenerative disease. The analysis successfully recovered known TRE associations with neurodegenerative diseases, which validates the pipeline’s ability to detect meaningful associations. By leveraging biobank-scale sample sizes across UKB and AoU, we can substantially increase power. The use of composite neurodegenerative phenotypes enables detection of pleiotropic effects across the neurodegenerative disease spectrum and can also increase power. The phenotypic breadth observed among carriers of established TRE loci further illustrates that known pathogenic expansions present with greater clinical variability than traditionally captured. Together, these findings implicate several candidate risk genes and loci for follow-up, including significant associations at *DAPK1*, *ANK3*, and *RPL14*, and suggestive signals at additional loci with biologically plausible roles in neurodegeneration.

## Supplemental Information

Supplemental Information includes 17 figures and 25 tables.

## Data Availability

UK Biobank data are available to approved researchers through the UK Biobank Resource. All of Us data used in this study are from the All of Us Research Program's Controlled Tier Dataset v9, available to authorized users on the Researcher Workbench. GTEx v8 expression and covariate data used in this study are available through the GTEx Portal (open-access tier); individual-level GTEx WGS genotypes are not hosted on the Portal and require separate controlled access via dbGaP (accession phs000424). MESA whole-genome sequencing data used for tandem repeat genotyping and RNA-seq expression data are available through dbGaP (accession phs001416). AMP-PD data are available through the AMP-PD platform. Individual-level data generated in this study cannot be shared publicly due to the data access policies of the UK Biobank and All of Us Research Program. Summary-level data not included in the manuscript or supplementary materials are available from the corresponding author upon reasonable request. Analysis scripts are available at https://github.com/gabrielle-altman/TRE_neuroDegen.

## Acknowledgments

This research has been conducted using the UKB Resource under application number 82094. We gratefully acknowledge All of Us participants for their contributions, without whom this research would not have been possible. We also thank the National Institutes of Health’s All of Us Research Program for making available the participant data examined in this study. An exception to the All of Us Data and Statistics Dissemination Policy, granted by the All of Us Resource Access Board, permitted the reporting of group sizes of fewer than 20 participants. The Genotype-Tissue Expression (GTEx) Project was supported by the Common Fund of the Office of the Director of the National Institutes of Health, and by NCI, NHGRI, NHLBI, NIDA, NIMH, and NINDS. The data used for the analyses described in this manuscript were obtained from: the GTEx Portal and dbGaP accession number phs000424.

Molecular data for the Trans-Omics in Precision Medicine (TOPMed) program was supported by the National Heart, Lung and Blood Institute (NHLBI). Genome sequencing for “NHLBI TOPMed: MESA” (phs001416) was performed at Broad Institute Genomics Platform (HHSN268201600034I). Genome sequencing for “NHLBI TOPMed: MESA AA_CAC” (phs001416) was performed at Broad Institute Genomics Platform (HHSN268201500014C). RNASeq for “NHLBI TOPMed: MESA” (phs001416) was performed at Broad Institute Genomics Platform (HHSN268201600034I) and Northwest Genomics Center (HHSN268201600032I). Core support including centralized genomic read mapping and genotype calling, along with variant quality metrics and filtering were provided by the TOPMed Informatics Research Center (3R01HL-117626-02S1; contract HHSN268201800002I). Core support including phenotype harmonization, data management, sample-identity QC, and general program coordination were provided by the TOPMed Data Coordinating Center (R01HL-120393; U01HL-120393; contract HHSN268201800001I). We gratefully acknowledge the studies and participants who provided biological samples and data for TOPMed.

MESA and the MESA SHARe projects are conducted and supported by the National Heart, Lung, and Blood Institute (NHLBI) in collaboration with MESA investigators. Support for MESA is provided by contracts 75N92020D00001, HHSN268201500003I, N01-HC-95159, 75N92020D00005, N01-HC-95160, 75N92020D00002, N01-HC-95161, 75N92020D00003, N01-HC-95162, 75N92020D00006, N01-HC-95163, 75N92020D00004, N01-HC-95164, 75N92020D00007, N01-HC-95165, N01-HC-95166, N01-HC-95167, N01-HC-95168, N01-HC-95169, UL1-TR-000040, UL1-TR-001079, and UL1-TR-001420, UL1TR001881, DK063491, and R01HL105756. Funding for SHARe genotyping was provided by NHLBI Contract N02-HL- 64278. Genotyping was performed at Affymetrix (Santa Clara, California, USA) and the Broad Institute of Harvard and MIT (Boston, Massachusetts, USA) using the Affymetrix Genome-Wide Human SNP Array 6.0. MESA Family is conducted and supported by the National Heart, Lung, and Blood Institute (NHLBI) in collaboration with MESA investigators. Support is provided by grants and contracts R01HL071051, R01HL071205, R01HL071250, R01HL071251, R01HL071258, R01HL071259, UL1TR001881, DK063491, and by the National Center for Research Resources, Grant UL1RR033176. The authors thank the other investigators, the staff, and the participants of the MESA study for their valuable contributions. A full list of participating MESA investigators and institutes can be found at http://www.mesa-nhlbi.org.

Data used in the preparation of this article were obtained from the Accelerating Medicine Partnership® (AMP®) Parkinson’s Disease (AMP PD) and Parkinson’s Disease & Related Disorders (AMP PDRD) Knowledge Platform. For up-to-date information on the study, visit https://www.amp-pdrd.org. The AMP® PD program is a public-private partnership managed by the Foundation for the National Institutes of Health and funded by the National Institute of Neurological Disorders and Stroke (NINDS) in partnership with the Food and Drug Administration (FDA), National Institute on Aging (NIA), Aligning Science Across Parkinson’s (ASAP) initiative; Celgene Corporation, a subsidiary of Bristol-Myers Squibb Company; GlaxoSmithKline plc (GSK); The Michael J. Fox Foundation for Parkinson’s Research (MJFF); AbbVie Inc.; Pfizer Inc.; Sanofi US Services Inc.; and Verily Life Sciences LLC. ACCELERATING MEDICINES PARTNERSHIP and AMP are registered service marks of the U.S. Department of Health and Human Services.

Clinical data and biosamples used in preparation of this article were obtained from the (i) Michael J. Fox Foundation for Parkinson’s Research (MJFF) and National Institutes of Neurological Disorders and Stroke (NINDS) BioFIND study, (ii) NINDS Parkinson’s Disease Biomarkers Program (PDBP), and (iii) MJFF Parkinson’s Progression Markers Initiative (PPMI).

BioFIND is sponsored by The Michael J. Fox Foundation for Parkinson’s Research (MJFF) with support from the National Institute for Neurological Disorders and Stroke (NINDS). The BioFIND Investigators have not participated in reviewing the data analysis or content of the manuscript. For up-to-date information on the study, visit www.michaeljfox.org/biofind. PPMI is sponsored by The Michael J. Fox Foundation for Parkinson’s Research and supported by a consortium of scientific partners: https://www.ppmi-info.org/about-ppmi/who-we-are/study-sponsors. The PPMI investigators have not participated in reviewing the data analysis or content of the manuscript. For up-to-date information on the study, visit www.ppmi-info.org. The Parkinson’s Disease Biomarker Program (PDBP) consortium is supported by the National Institute of Neurological Disorders and Stroke (NINDS) at the National Institutes of Health. A full list of PDBP investigators can be found at https://pdbp.ninds.nih.gov/policy. The PDBP investigators have not participated in reviewing the data analysis or content of the manuscript.

This work was supported by NIH grant AG075051 to A.J.S. and through the computational resources and staff expertise provided by Scientific Computing at the Icahn School of Medicine at Mount Sinai, supported by the Clinical and Translational Science Awards (CTSA) grant UL1TR004419 from the National Center for Advancing Translational Sciences and by the Office of Research Infrastructure of the NIH under award number S10OD026880. The content is solely the responsibility of the authors and does not necessarily represent the official views of the National Institutes of Health. The funders had no role in study design, data collection and analysis, decision to publish, or preparation of the manuscript.

## Data and code availability

UK Biobank data are available to approved researchers through the UK Biobank Resource. All of Us data used in this study are from the All of Us Research Program’s Controlled Tier Dataset v9, available to authorized users on the Researcher Workbench. GTEx v8 expression and covariate data used in this study are available through the GTEx Portal (open-access tier); individual-level GTEx WGS genotypes are not hosted on the Portal and require separate controlled access via dbGaP (accession phs000424). MESA whole-genome sequencing data used for tandem repeat genotyping and RNA-seq expression data are available through dbGaP (accession phs001416). AMP-PD data are available through the AMP-PD platform. Individual-level data generated in this study cannot be shared publicly due to the data access policies of the UK Biobank and All of Us Research Program. Summary-level data not included in the manuscript or supplementary materials are available from the corresponding author upon reasonable request. Analysis scripts are available at https://github.com/gabrielle-altman/TRE_neuroDegen.

## Author contributions

Conceptualization and study design, G.N.A, A.M.T, and A.J.S.; data curation and genotyping, G.N.A., B.J., P.G., M.S., C.A.M., W.L., S.K., and A.M.T.; phenotype curation, G.N.A., B.J., and W.L.; formal analysis, G.N.A.; software and pipeline development, G.N.A., B.J., P.G., M.S., C.A.M., W.L., S.K., and A.M.T.; visualization, G.N.A.; supervision, A.M.T. and A.J.S.; funding acquisition, A.J.S.; writing – original draft, G.N.A.; writing – review & editing, all authors. All authors have reviewed and approved the final version of the manuscript.

## Declaration of generative AI and AI-assisted technologies in the writing process

During the preparation of this work, the authors used Claude (Anthropic) and Grammarly in order to improve grammar, clarity, and readability. After using these tools, the authors reviewed and edited the content as needed and take full responsibility for the content of the publication.

## Web Resources

All of Us Researcher Workbench, https://workbench.verily.com

UK Biobank, https://www.ukbiobank.ac.uk

ExpansionHunter, https://github.com/Illumina/ExpansionHunter

TRGT, https://github.com/pacificBiosciences/trgt/

REViewer, https://github.com/illumina/REViewer

REGENIE, https://rgcgithub.github.io/regenie/

METAL, https://github.com/statgen/METAL

Classifier, https://zenodo.org/doi/10.5281/zenodo.10821643

Global ancestry assignment, https://zenodo.org/records/10820995

CAVIAR, http://genetics.cs.ucla.edu/caviar/

brglm, https://cran.r-project.org/web/packages/brglm/index.html

PLINK 1.9, www.cog-genomics.org/plink/1.9/

PLINK 2.0, www.cog-genomics.org/plink/2.0/

ANNOVAR, https://annovar.openbioinformatics.org

GTEx Portal, https://gtexportal.org

AMP-PD, https://amp-pd.org

MESA/TOPMed, https://topmed.nhlbi.nih.gov/project-group/mesa

Scripts, https://github.com/gabrielle-altman/TRE_neuroDegen

