## Supplemental Figures for "Tandem repeat expansions in *DAPK1*, *ANK3*, and *RPL14* are associated with diverse neurodegenerative diseases"

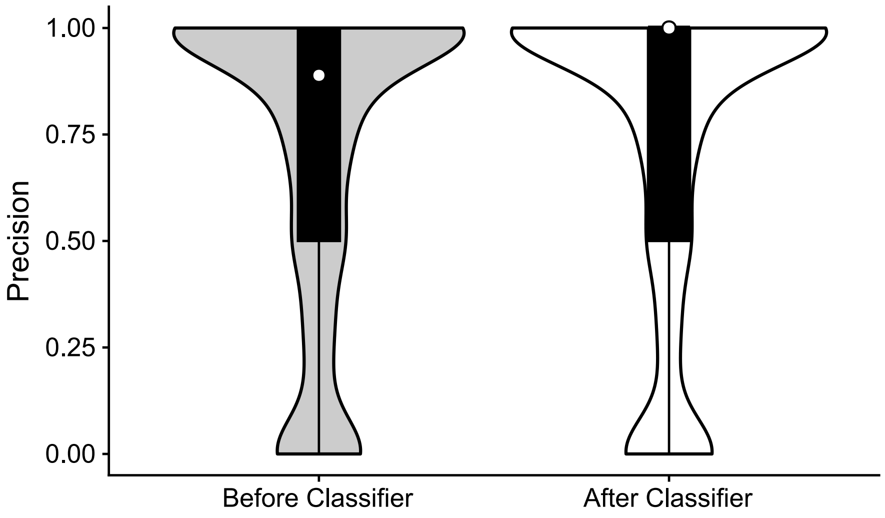


**Figure S1. EHv5 genotyping precision at genic and promoter tandem repeat loci, before and after classifier-based filtering.** Precision was calculated per locus as the proportion of ExpansionHunter v5 (EHv5) expansion calls from short-read sequencing data that were concordant with TRGT expansion calls from long-read sequencing data, restricted to loci annotated as genic or promoters. Violin plots show the distribution of per-locus precision across all qualifying loci "Before Classifier" and "After Classifier" filtering; overlaid black boxplots denote the interquartile range and median, and white circles mark the median precision for each group. Precision values are bounded between 0 and 1. The median precision was 0.889 before the classifier and was 1 after the classifier.

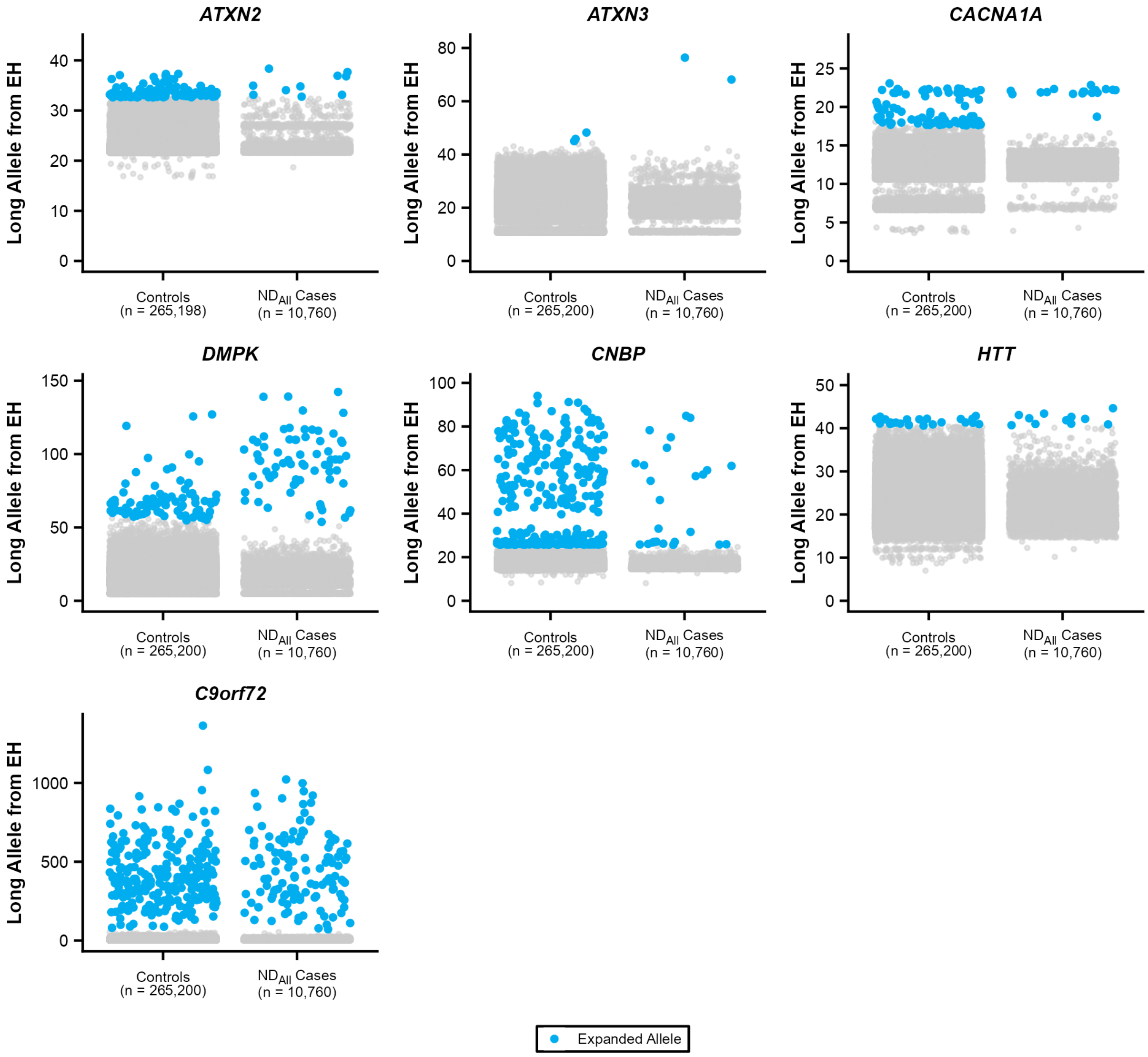


**Figure S2. Long allele size at known pathogenic repeat expansion loci associated with ND$_{All}$ in UK Biobank.** Each panel shows one known pathogenic repeat locus (labeled by gene) nominally associated with the full neurodegenerative disease cohort (ND$_{All}$) versus controls, plotted as individual-level jitter plots as in Figure 4 (grey points, non-expanded alleles; blue points, expanded alleles).

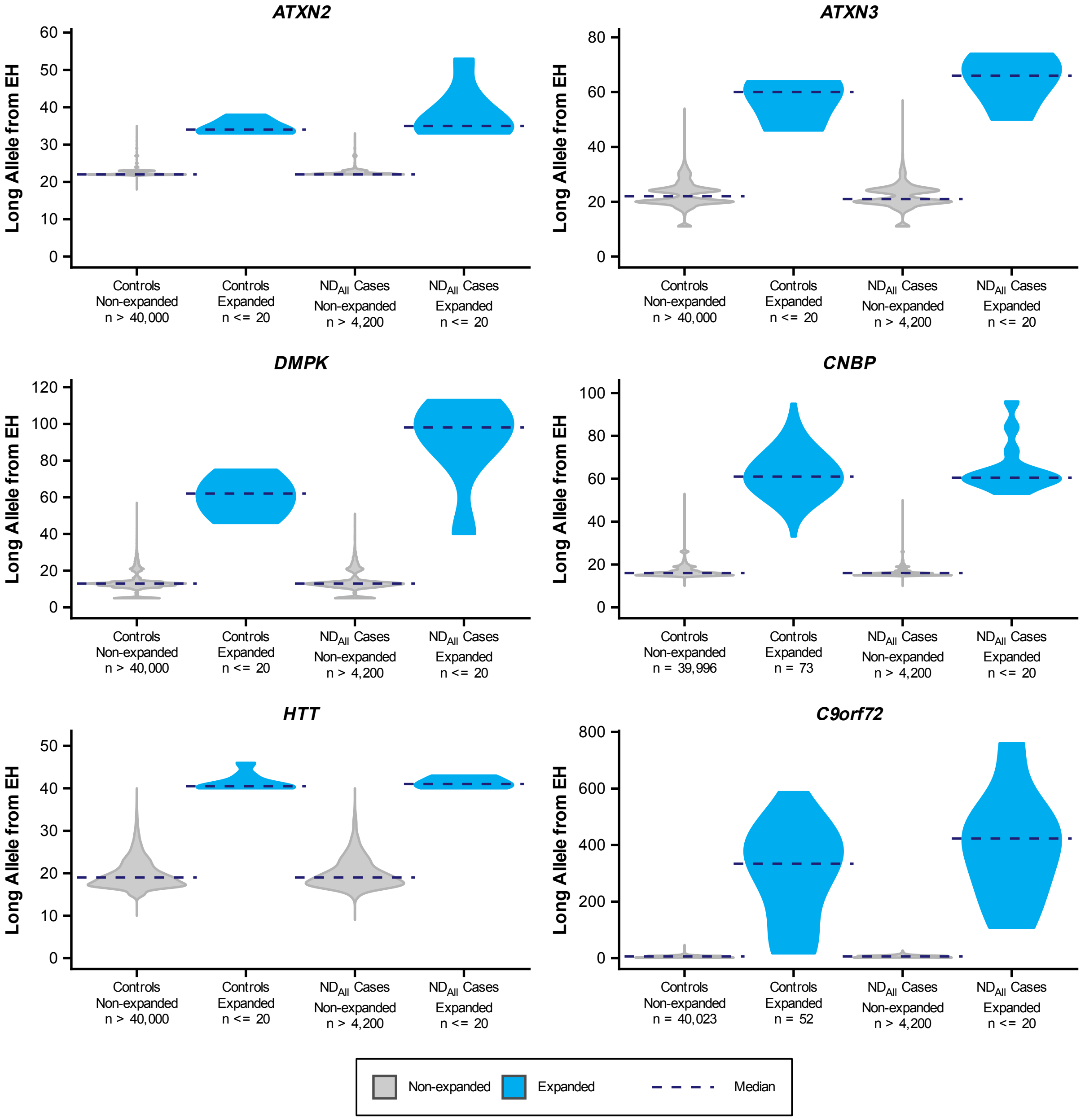


**Figure S3. Long allele size at known pathogenic repeat expansion loci associated with ND$_{All}$ in All of Us.** Each panel shows one known pathogenic repeat locus (labeled by gene) nominally associated with ND$_{All}$ versus controls, plotted as violin distributions split by case/control and expansion status as in Figure 4 (grey, non-expanded; blue, expanded; dashed navy line, group median). As in Figure 4, group sizes and near-uniform distributions are obscured as needed to comply with All of Us data use policies, without altering ORs or p-values.

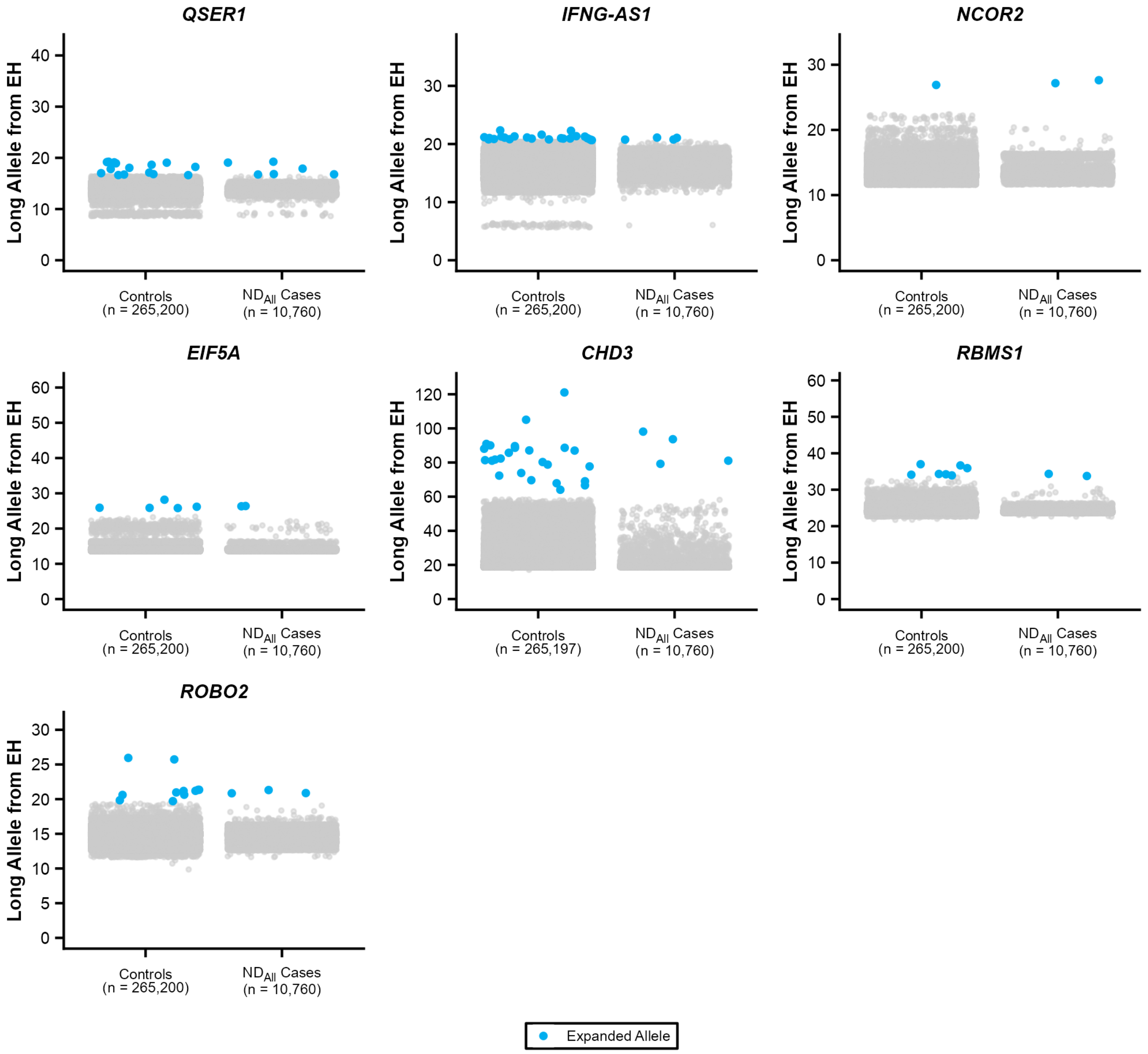


**Figure S4. Long allele size at candidate repeat expansion loci associated with ND$_{All}$ in UK Biobank.** As in Figure S2, but for non-pathogenic-annotated candidate loci nominally associated with ND$_{All}$. Panels exclude loci already shown in Figure 4.

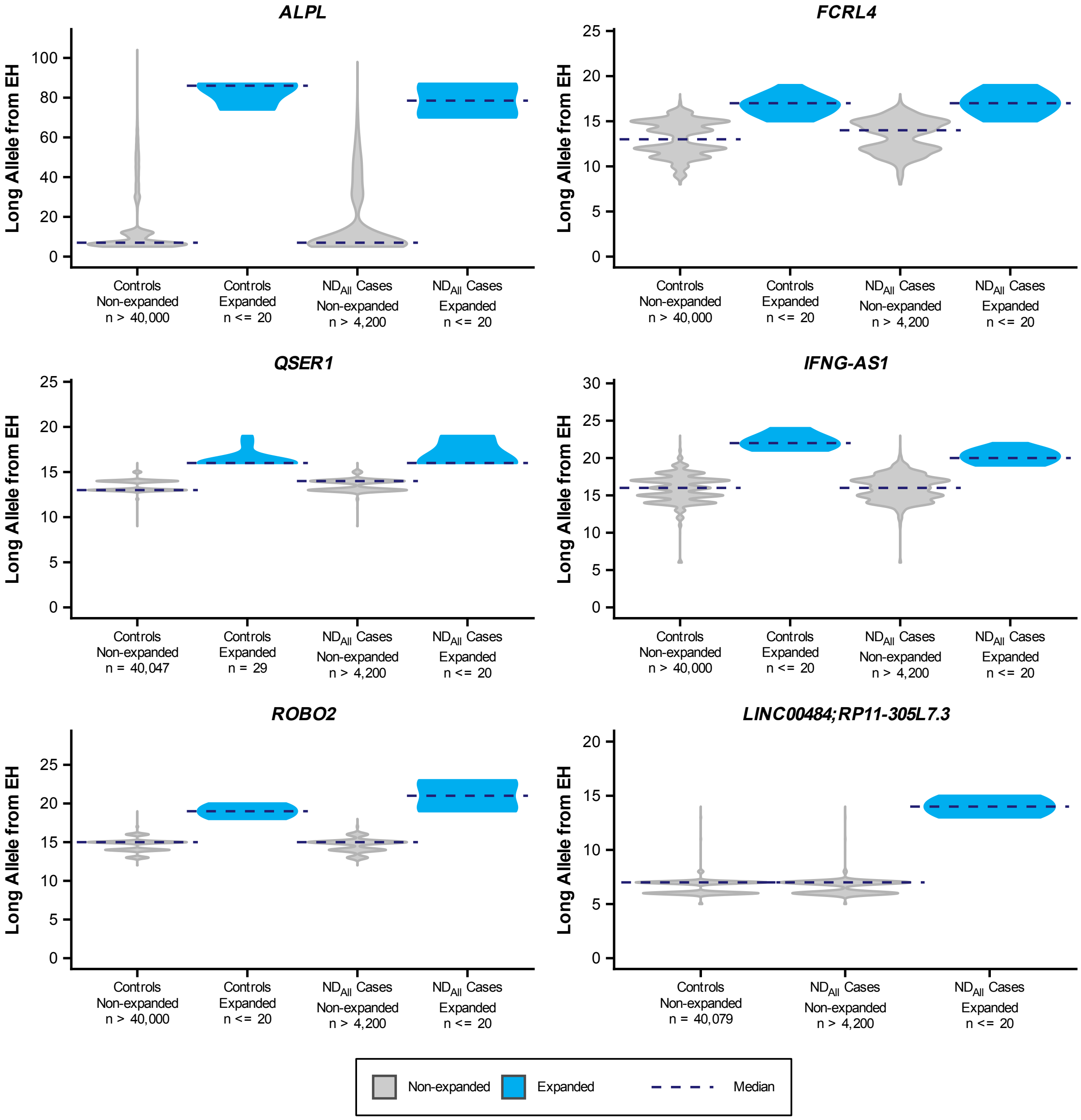


**Figure S5. Long allele size at candidate repeat expansion loci associated with ND$_{All}$ in All of Us.** As in Figure S3, but for candidate loci associated with ND$_{All}$. Panels exclude loci already shown in Figure 4.

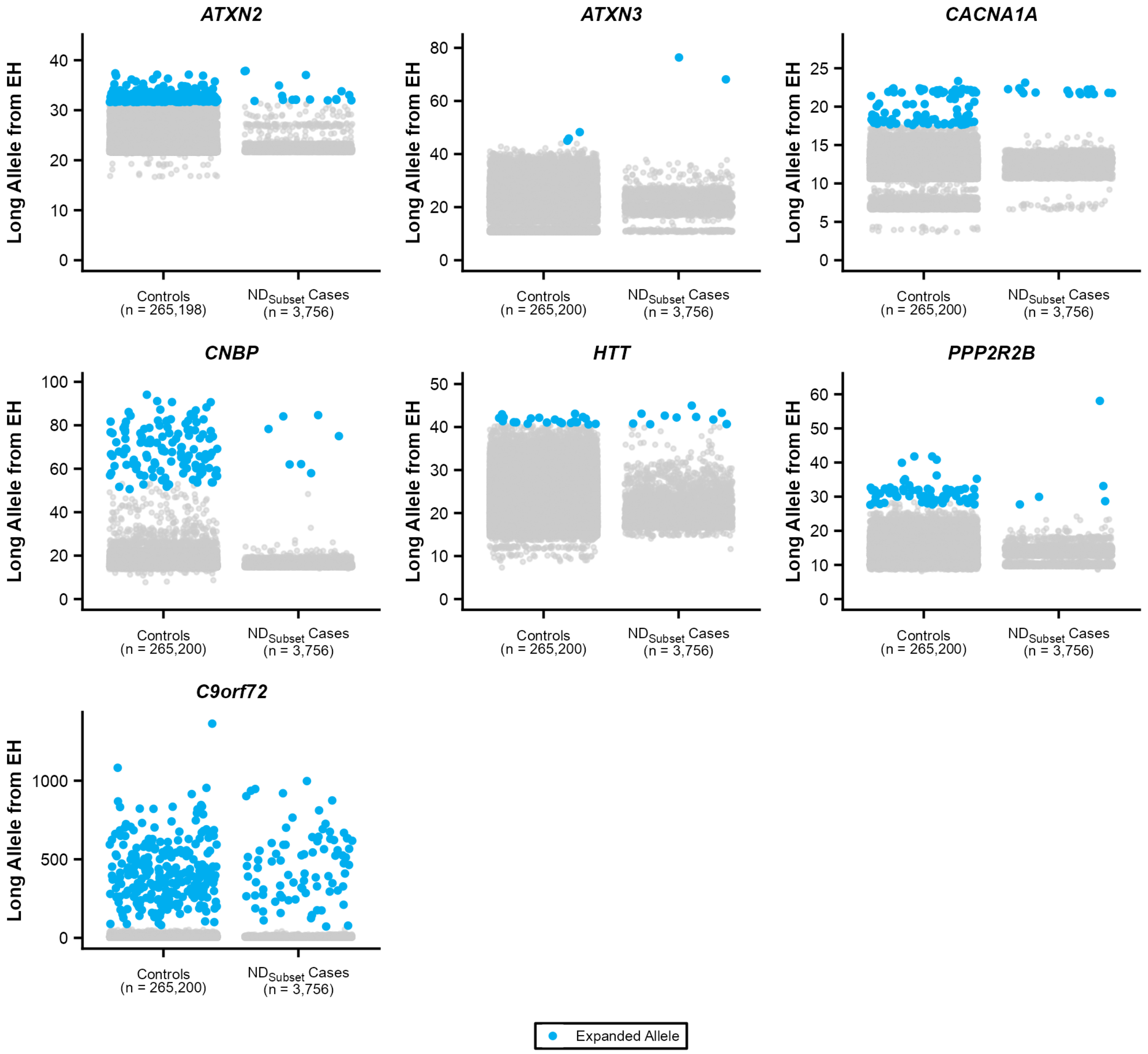


**Figure S6. Long allele size at known pathogenic repeat expansion loci associated with ND$_{Subset}$ in UK Biobank.** As in Figure S2, but for the neurodegenerative-disease cohort excluding Parkinson's and Alzheimer's disease (ND$_{Subset}$). Panels exclude loci already shown in Figure 4.

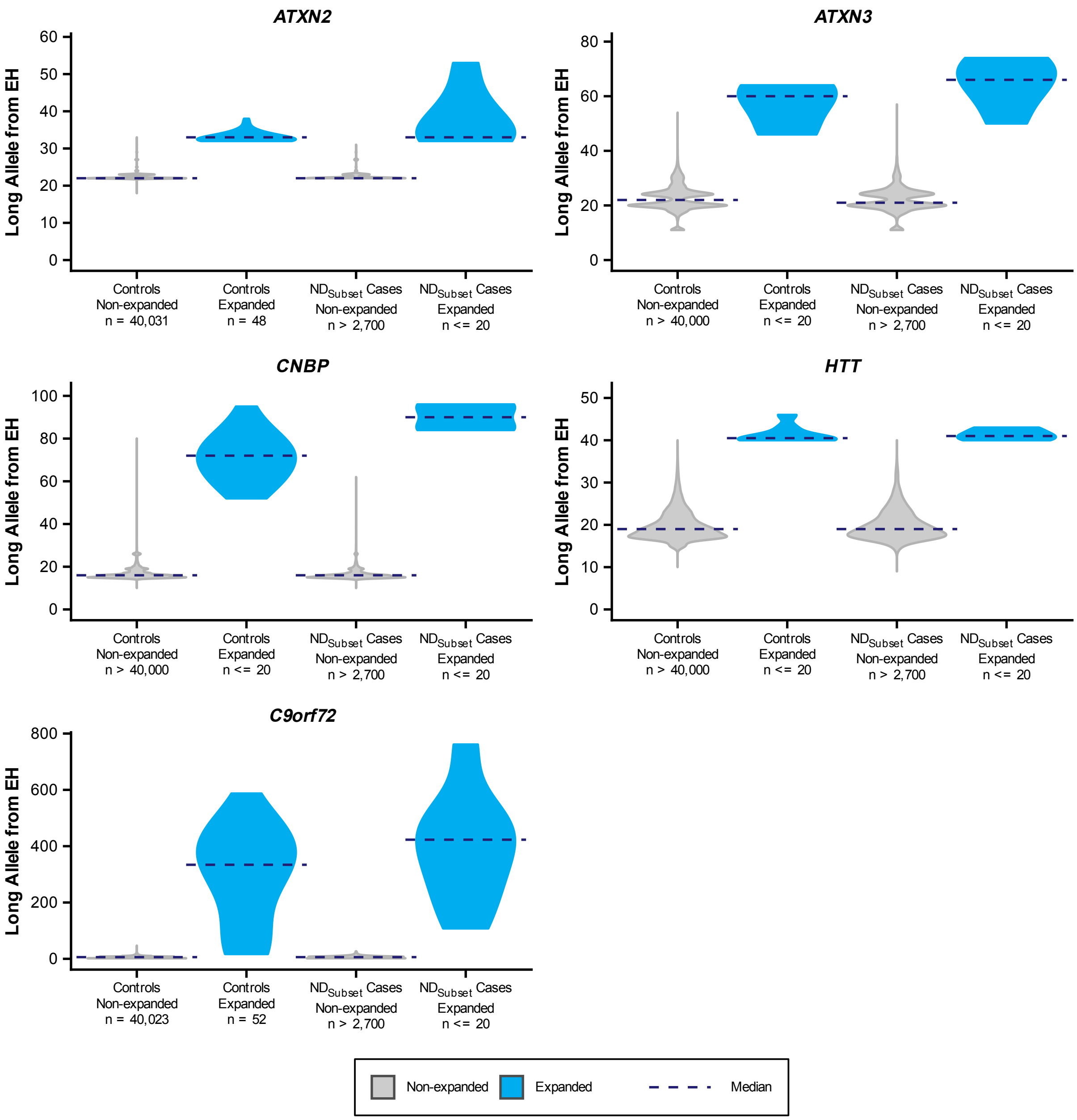


**Figure S7. Long allele size at known pathogenic repeat expansion loci associated with ND$_{Subset}$ in All of Us.** As in Figure S3, but for ND$_{Subset}$. Panels exclude loci already shown in Figure 4.

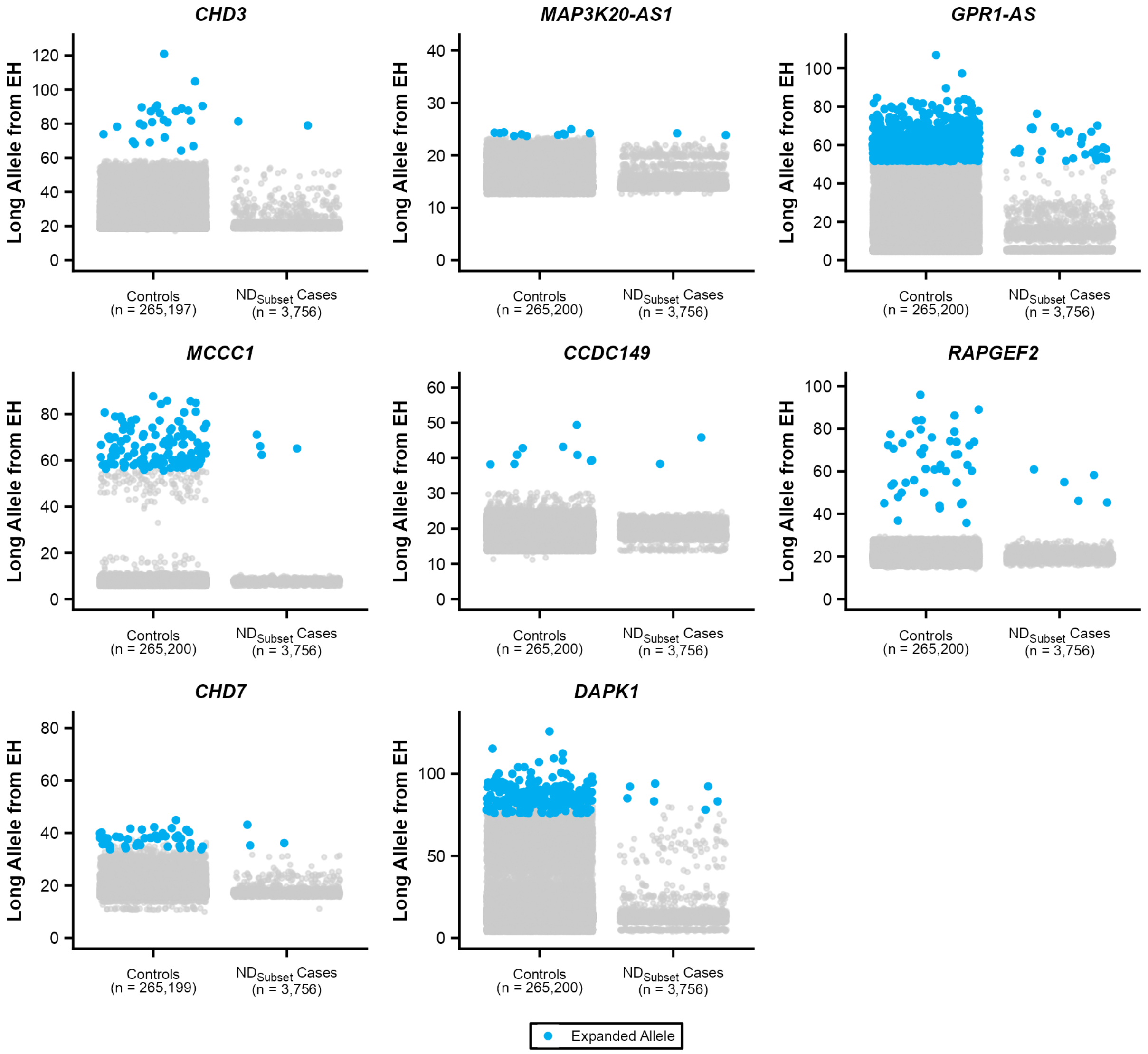


**Figure S8. Long allele size at candidate repeat expansion loci associated with ND$_{Subset}$ in UK Biobank.** As in Figure S4, but for ND$_{Subset}$. Panels exclude loci already shown in Figure 4.

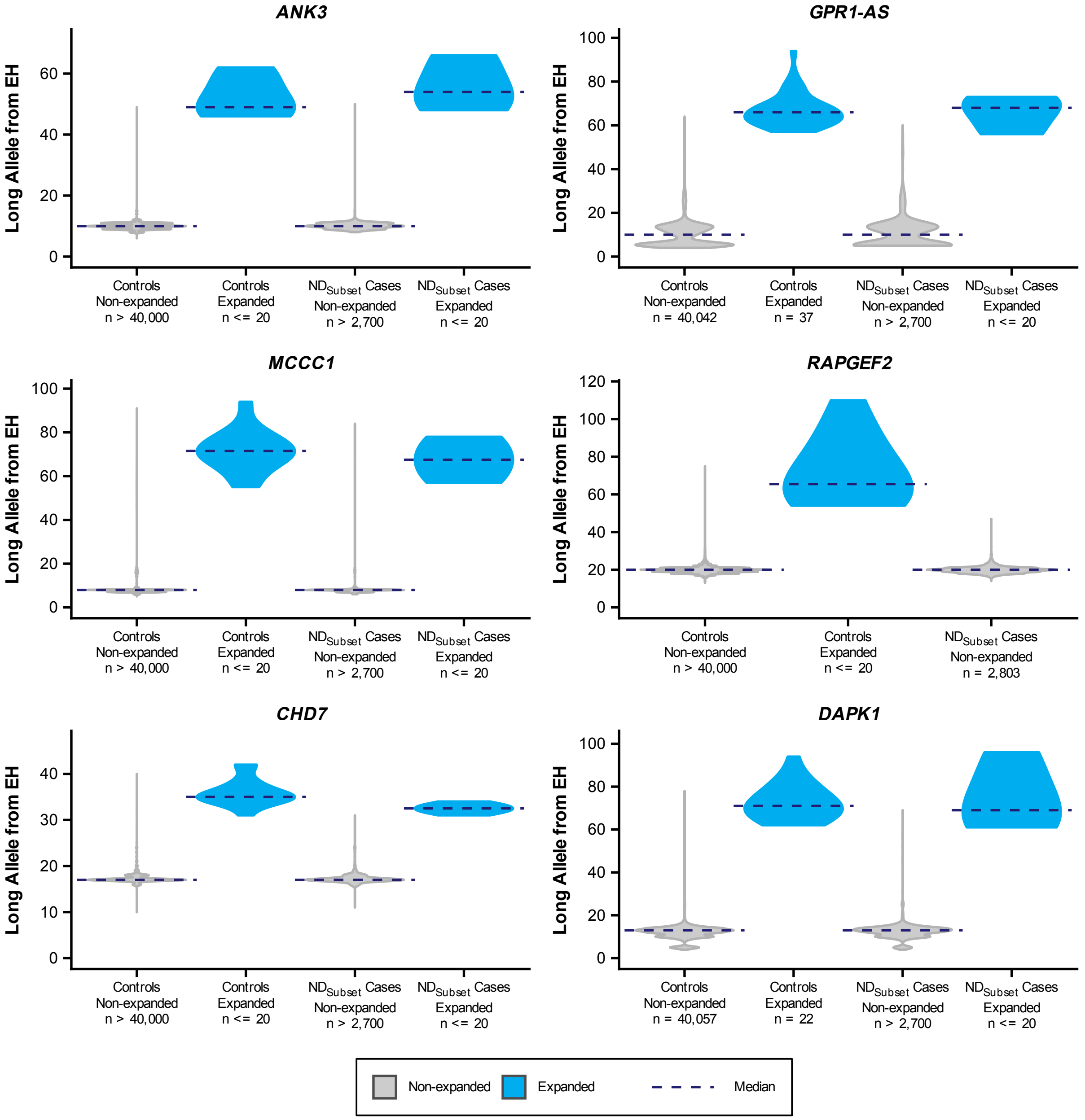


**Figure S9. Long allele size at candidate repeat expansion loci associated with ND$_{Subset}$ in All of Us.** As in Figure S5, but for ND$_{Subset}$. Panels exclude loci already shown in Figure 4.

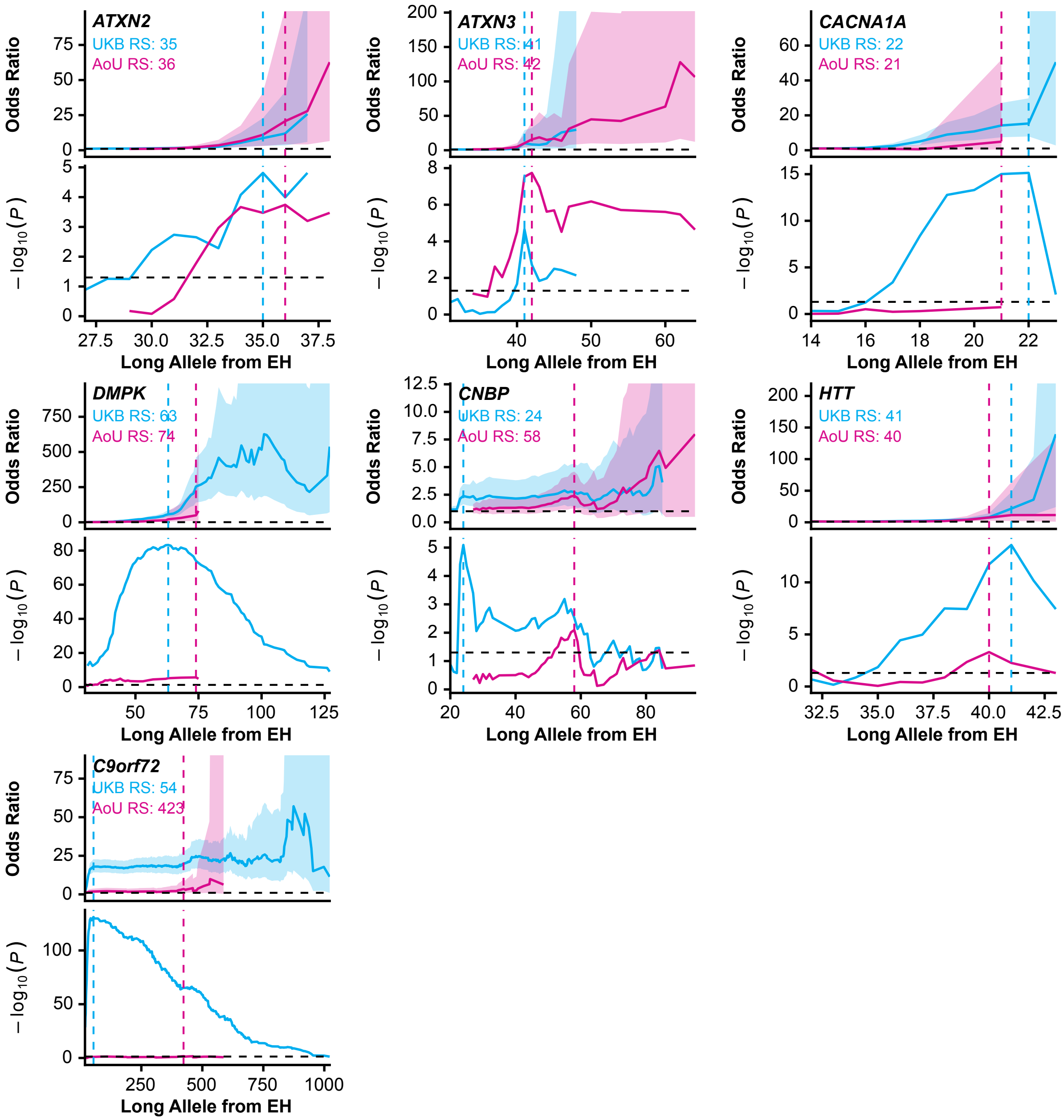


**Figure S10. Odds ratio and significance across candidate repeat-length thresholds for known pathogenic loci associated with $ND_{All}$, UK Biobank and All of Us.** Each locus (labeled by gene) is shown as a stacked two-row panel: odds ratio with 95% confidence interval for UK Biobank and for All of Us, each as a function of the long-allele repeat-length threshold used to call an expansion, and the corresponding $-\log_{10}(p)$ curves for both cohorts. Both plots show UKB and AoU data overlaid on a shared axis (blue: UKB; pink: AoU). Both rows share the same x-axis range. Dashed horizontal lines mark OR = 1 (top plot) and the p = 0.05 significance threshold (bottom plot); dashed vertical lines mark the repeat-length threshold giving the lowest p-value in each cohort (blue: UKB; pink: AoU).

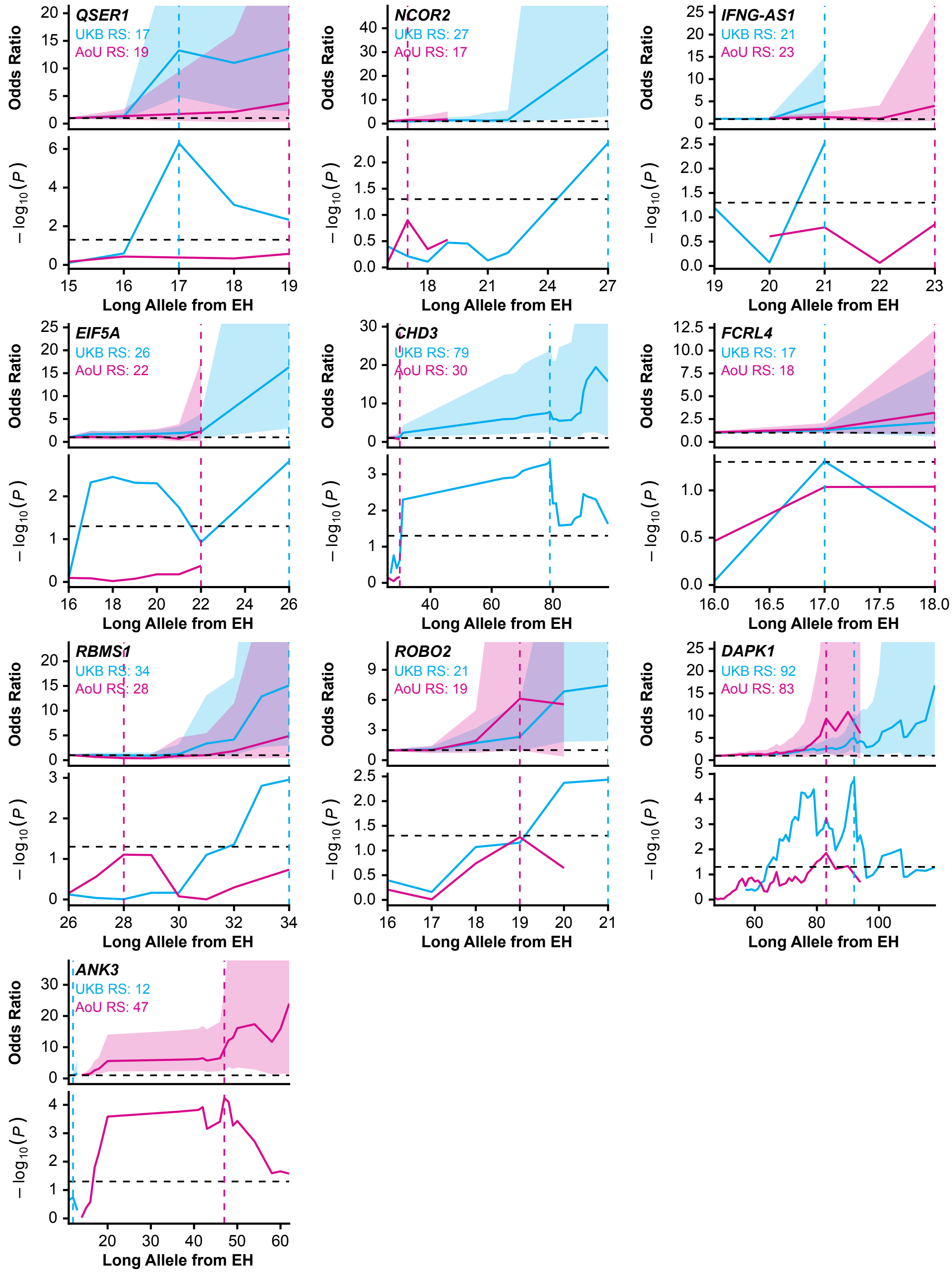


**Figure S11. Odds ratio and significance across candidate repeat-length thresholds for candidate loci associated with $ND_{All}$, UK Biobank and All of Us.** As in Figure S10, but for non-pathogenic-annotated candidate loci nominally associated with $ND_{All}$.

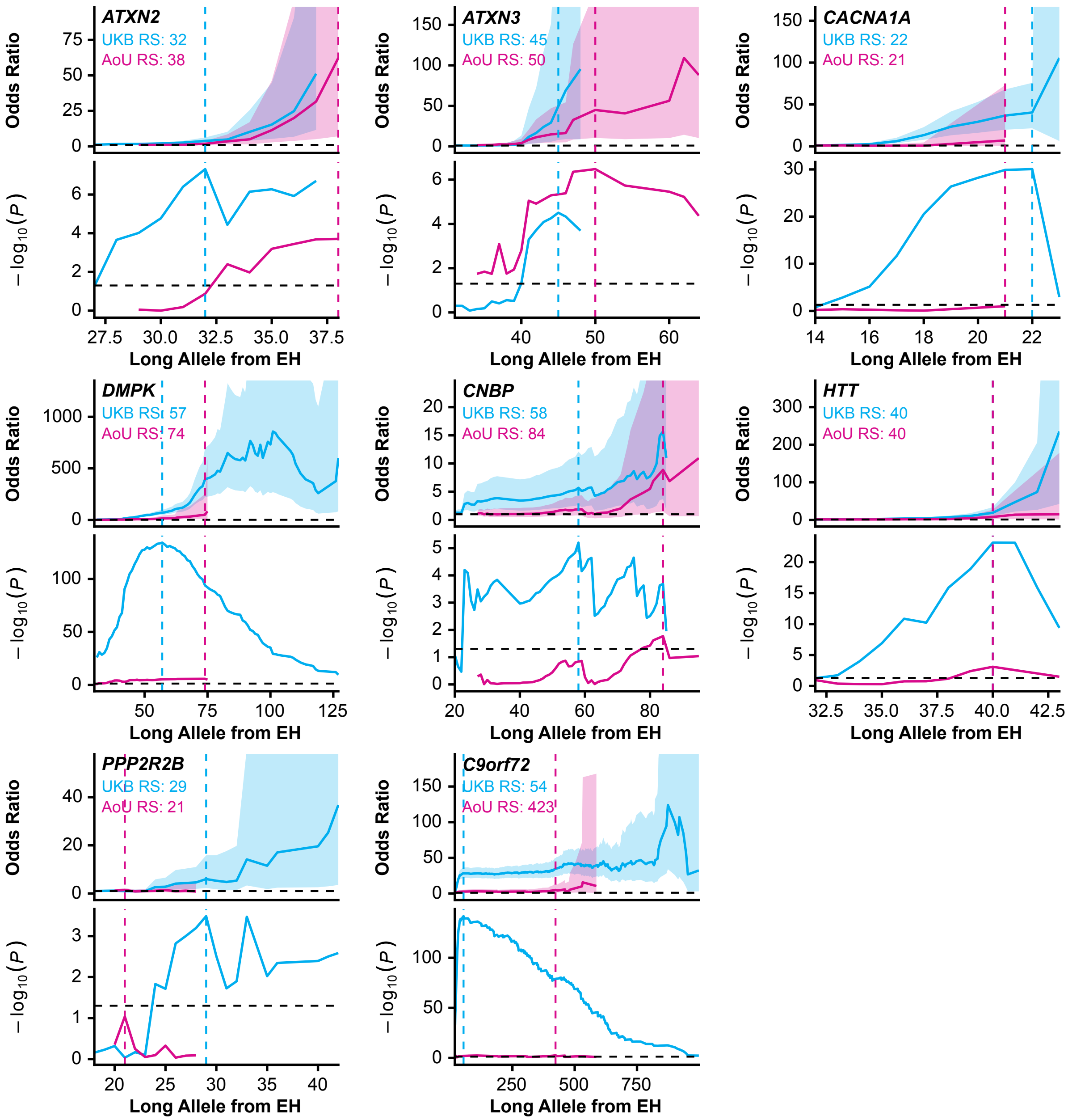


**Figure S12. Odds ratio and significance across candidate repeat-length thresholds for known pathogenic loci associated with $ND_{Subset}$, UK Biobank and All of Us.** As in Figure S10, but for the neurodegenerative-disease cohort excluding Parkinson's and Alzheimer's disease ($ND_{Subset}$).

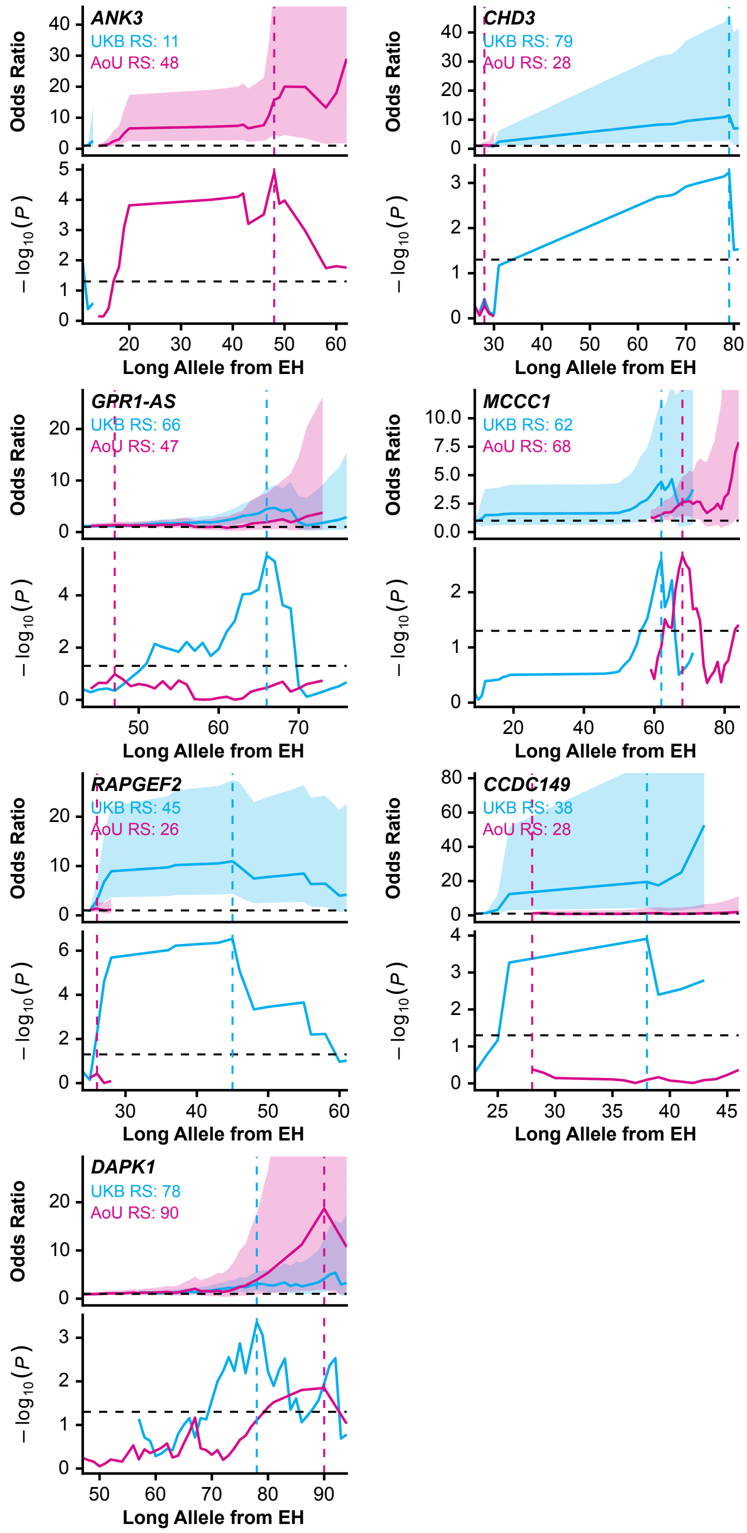


**Figure S13. Odds ratio and significance across candidate repeat-length thresholds for candidate loci associated with $ND_{Subset}$, UK Biobank and All of Us.** As in Figure S11, but for $ND_{Subset}$.

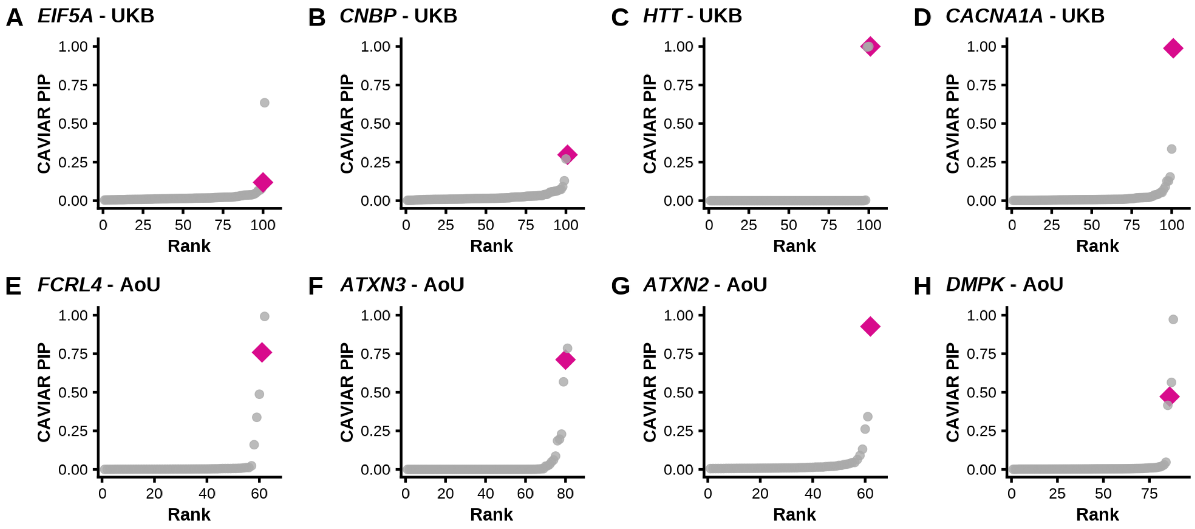


**Figure S14. CAVIAR fine-mapping posterior inclusion probabilities at eight candidate tandem repeat loci in UK Biobank and All of Us.** CAVIAR fine-mapping results at eight candidate loci: (A–D) UK Biobank: *EIF5A, CNBP, HTT, CACNA1A*; (E–H) All of Us: *FCRL4, ATXN3, ATXN2, DMPK*. Each panel ranks the top 100 candidate variants at that locus by CAVIAR posterior inclusion probability (PIP; y-axis) against rank (x-axis). Grey circles, SNVs; pink diamond, the short tandem repeat (STR) included as a candidate causal variant in the fine-mapping model. All tests were done with the $ND_{All}$ phenotype.

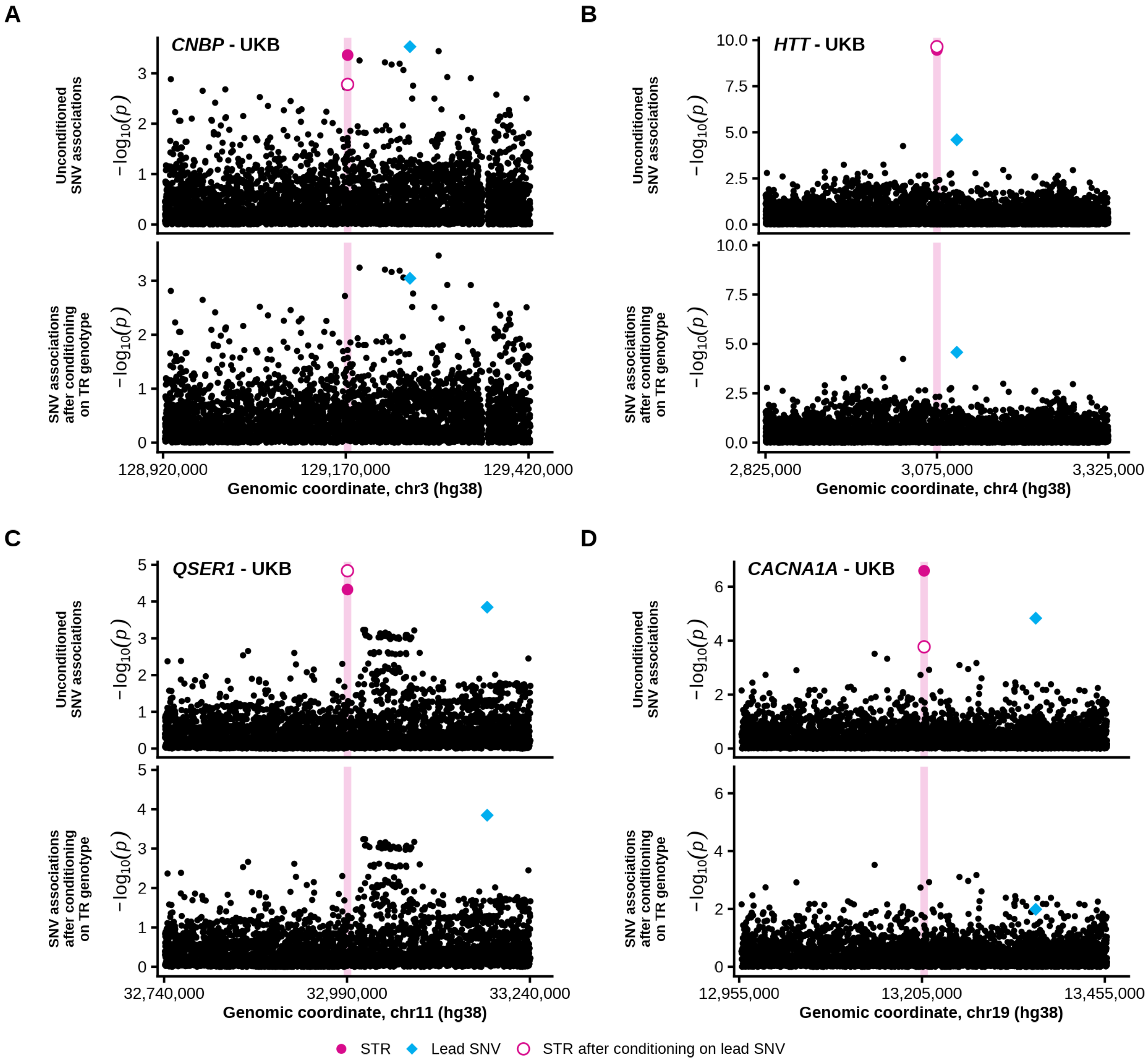


**Figure S15. Conditional association analysis of tandem repeat and lead single-nucleotide variant signals in UK Biobank.** Two-panel conditional association scans for four loci: *CNBP, HTT, QSER1, CACNA1A*. For each locus: top, unconditioned $-\log_{10}$(p) association across the genomic region (grey points, SNVs; filled pink circle, the STR's own unconditioned association; blue diamond, the lead SNV; open pink circle, the STR's association after conditioning on the lead SNV genotype, overlaid for comparison); bottom, the SNV association scan after conditioning on the STR's genotype (grey points, SNVs; blue diamond, the lead SNV), testing whether the SNV signal persists once the STR's contribution is removed. The shaded pink vertical line marks the STR's genomic position in both panels; x-axis, genomic coordinate (hg38); y-axis, $-\log_{10}$(p). All tests were done with the $ND_{All}$ phenotype.

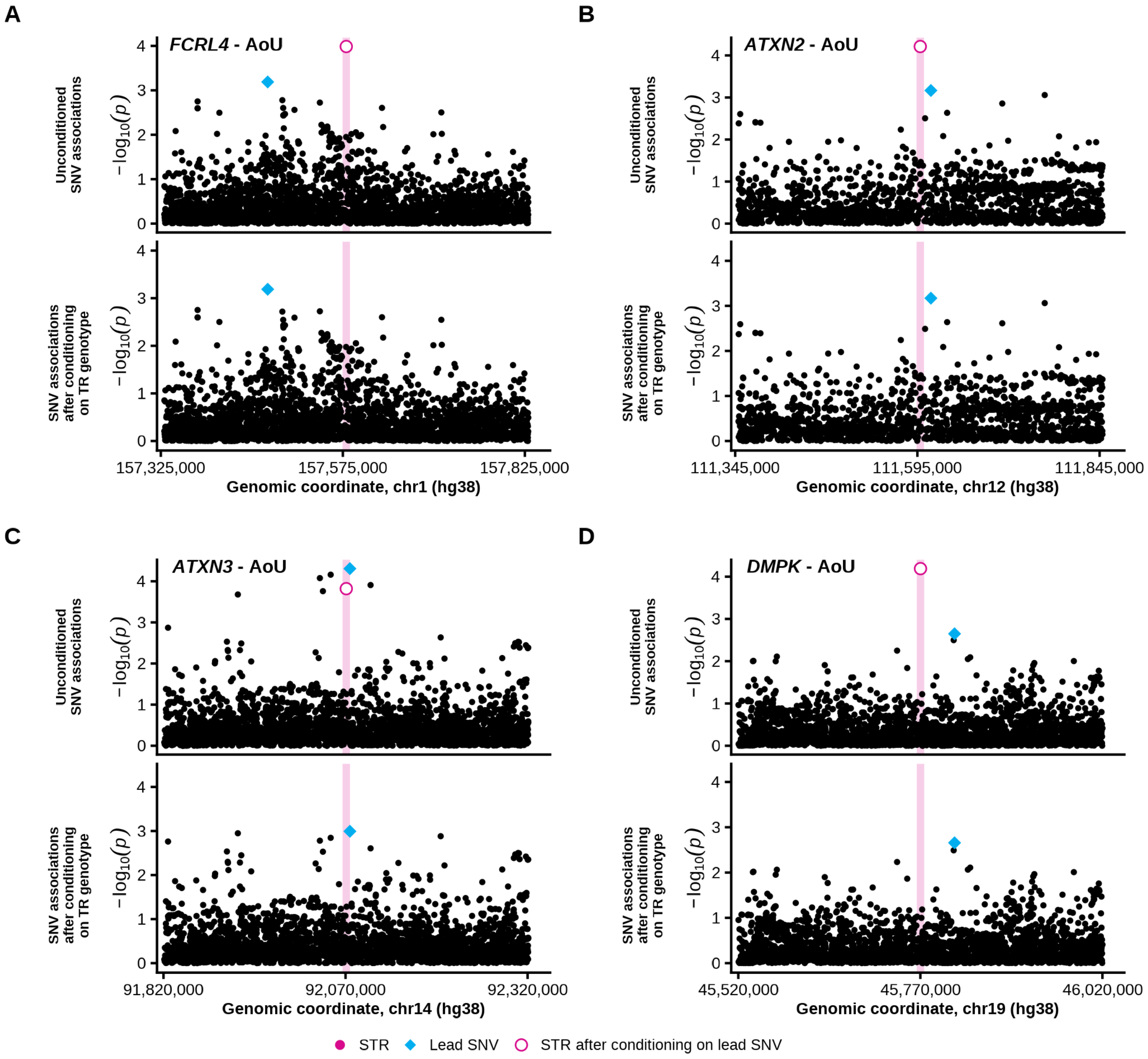


**Figure S16. Conditional association analysis of tandem repeat and lead single-nucleotide variant signals in All of Us.** As in Figure S15, but for four loci in All of Us: *FCRL4, ATXN2, ATXN3, DMPK*. All tests were done with the $ND_{All}$ phenotype.

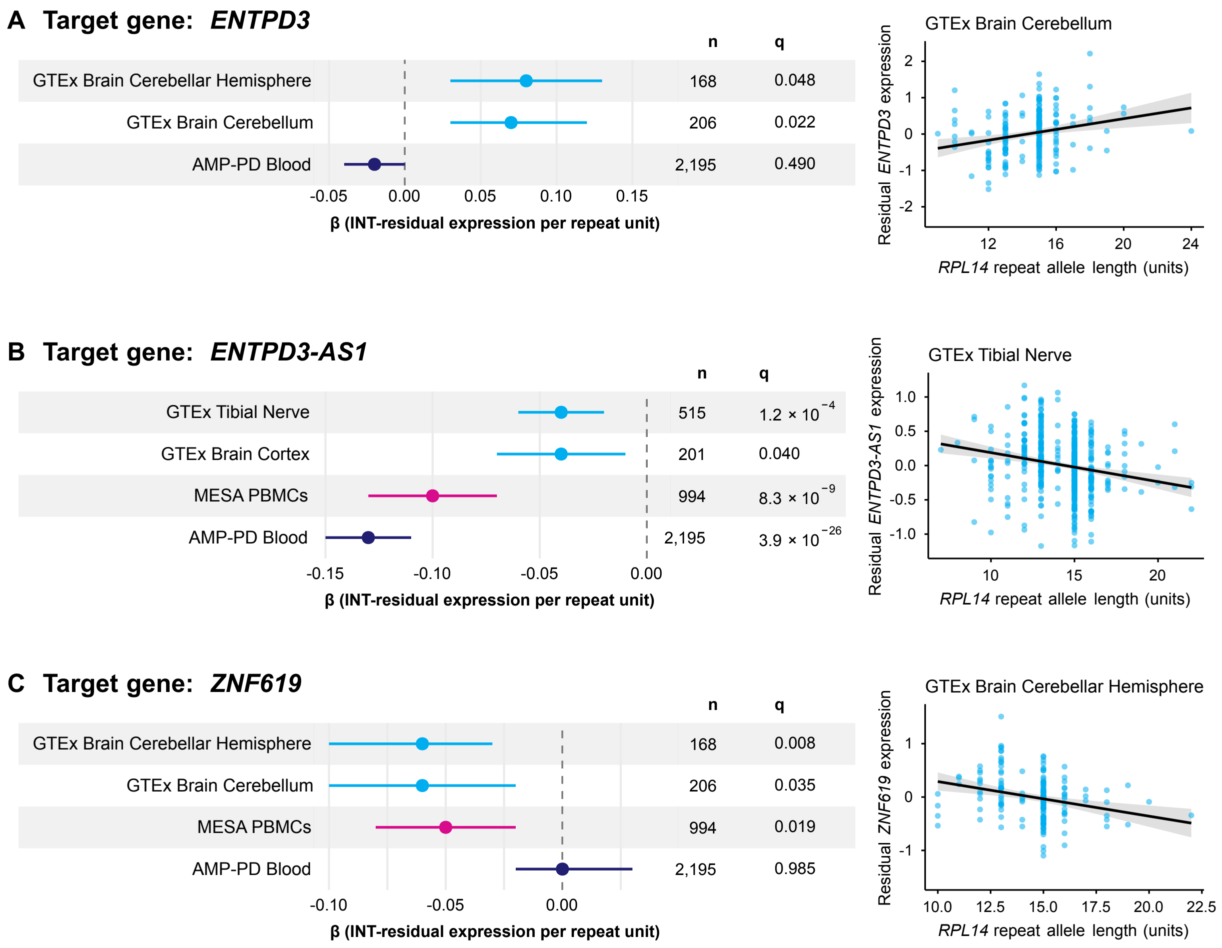


**Figure S17. The *RPL14* repeat locus is associated with expression of neighboring genes *ENTPD3, ENTPD3-AS1*, and *ZNF619*.** For each gene, forest plots (left) show the effect (β, INT-residualized expression per repeat unit) of the *RPL14* repeat locus (chr3:40,462,029–40,462,059) on expression across datasets, with 95% CI, sample size (n), and FDR-corrected q-value. Scatter plots (right) show residual expression against *RPL14* repeat allele length in a representative tissue, with the regression line and 95% CI shown. (A) *ENTPD3* expression in GTEx Brain Cerebellar Hemisphere, GTEx Brain Cerebellum, and AMP-PD Blood; scatter plot shows GTEx Brain Cerebellum. (B) *ENTPD3-AS1* expression in GTEx Tibial Nerve, GTEx Brain Cortex, MESA PBMCs, and AMP-PD Blood; scatter plot shows GTEx Tibial Nerve. (C) *ZNF619* expression in GTEx Brain Cerebellar Hemisphere, GTEx Brain Cerebellum, MESA PBMCs, and AMP-PD Blood; scatter plot shows GTEx Brain Cerebellar Hemisphere.
